# PULSE PRESSURE IMPAIRS COGNITION VIA WHITE MATTER DISRUPTION

**DOI:** 10.1101/2024.12.20.24319319

**Authors:** Deborah L. O. King, Richard N. Henson, Marta Correia, James B. Rowe, Kamen A. Tsvetanov

**Author notes:** Correspondence should be addressed to Kamen Tsvetanov, Department of Clinical Neurosciences, Cambridge University, Cambridge CB2 0SZ, United Kingdom.

## Abstract

**BACKGROUND:** In older adults, elevated pulse pressure predicts cognitive decline, irrespective of overall blood pressure. It is proposed to compromise cerebrovascular integrity, potentially leading to brain damage, though the underlying mechanisms remain unclear. We hypothesized that pulse pressure affects cognition by disrupting white matter microstructure, and that it does so independently of other cardiovascular risk factors.

**METHODS:** Indices of pulse pressure, overall blood pressure and heart rate variability were estimated in a cross-sectional population-based cohort (n=708, aged 18-88 years). An indicator of white matter microstructure was derived from diffusion-weighted imaging, termed the “peak width of skeletonised mean diffusivity” (PSMD). Cognitive function was assessed using measures of processing speed.

**RESULTS:** In robust multiple linear regressions, pulse pressure significantly predicted PSMD. We also found that PSMD significantly predicted processing speed. Thus higher pulse pressure was associated with greater white matter disruption, and greater white matter disruption was associated with slower processing abilities.This motivated testing whether PSMD mediates the effects of pulse pressure on processing speed. We tested this using a number of structural equation models. PSMD significantly and substantially mediated the effect of pulse pressure on processing speed, over and above age and other cardiovascular factors. We then expanded the model to show that vascular-related changes in processing speed in turn drive changes in higher cognitive functions.

**CONCLUSIONS:** High pulse pressure disrupts microstructural integrity of white matter in the brain, leading to slower processing speed. We propose that better manament of pulse pressure could help to preserve white matter integrity and reduce cognitive decline in later life.

## INTRODUCTION

With increasing, arteries lose elasticity and stiffen, resulting in an elevation of pulse pressure, the difference between systolic and diastolic blood pressure. The increased pulse pressure penetrates deeper into the cerebral microcirculation, where it is hypothesized to induce microvascular damage. This intitates a cascade of events^1^, including impaired cerebral blood flow and subsequent hypoxia. White matter is particularly vulnerable to hypoxia^2^, because arterioles are widely spaced, narrow and long, making them less able to maintain perfusion than grey matter^3–6^. Hypoxia also impairs the ability of astrocytes and oligodendrocytes to repair white matter myelin. White matter ageing is associated with demyelination and cell death, and can progress to macrostructural changes. Such changes are detectable as the lesions and hyperintensities on magnetic resonance imaging scans, which are in turn associated with cognitive impairment, including but not limited to vascular dementia. Recent studies have associated the early and subtle changes in white matter microstructure with higher pulse pressure in middle and old age^7–10^. This raises the hypothesis that white matter microstructure may mediate specifically the relationship between pulse pressure and cognition, independently of other cardiovascular factors^11^.

Various measures of white matter microstructure can be derived from diffusion-weighted imaging (DWI), such as tensor-derived measures of mean diffusivity (MD) and fractional anisotropy (FA). DWI is based on the movement of water, where healthy tissues with rigid structures restrain water flow more than ageing tissues, in which axonal integrity degrades and perivascular spaces widen. DWI studies commonly average across white matter voxels globally, or within a tract of interest, to produce central tendency statistics, such as MD and FA. However, such averaging sacrifices sensitivity to tissue heterogeneity. An individual with a wide range of diffusion values across white matter voxels is more likely to have regions of focal damage, eg hyperintensities, somewhere in their brain, even if their mean MD/FA differs little from others.

An alternative approach in the face of heterogeneity is to quantify the ‘peak width’ distribution of skeletonised mean diffusivity (PSMD)^12^. PSMD represents the spread across the 90% voxels within the 5^th^ and 95^th^ percentiles of the distribution^12^. PSMD has been found to increase non-monotonically with age: slowly in young adulthood, and then accelerating after around 60 years of age^13^. This age profile suggests that PSMD may be an early marker of individual variations in ageing^14,15^. Furthermore, PSMD associates more strongly with markers of ischemic than neurodegenerative processes^12,16^ and is higher in individuals with cerebrovascular disease than healthy controls^15^, suggesting that it might be sensitive to cerebrovascular factors like pulse pressure. However, though PSMD has been associated with clinical status in diabetes, hypertension and smoking, it has not been related it to sub-clinical and continuous measures of vascular health – including pulse pressure – in healthy ageing^17,18^. It is important to understand what drives the increase in PSMD with age; and whether it is a mediating factor that links systemic vascular factors to cognitive ageing^19^.

There are strong empirical links between white matter health and cognition^20^. PSMD outperforms established markers of white matter injury in predicting cognitive performance^12,16,21,22^. The cognitive domain that relates to PSMD most consistently is processing speed^12,21–24^, as measured by a variety of speeded tasks. This may be because PSMD assesses microstructural integrity in the main white matter tracts, which contribute to the speed of information transmission across the brain. Processing speed is one of the first cognitive domains to decline with age, and it is thought to be the foundation for age-related differences in other cognitive domains^25–27^. Evidence for this comes from a previous study in the Cam-CAN cohort, which proposed and then tested a hierarchical “watershed” model^28^. This showed that individual differences in white matter predicted “downstream” differences in processing speed, which in turn predicted a substantial amount of variance in fluid intelligence^28^. Fluid intelligence is a crucial ability that underlies many cognitive tests, and declines steeply with age. Moreover, its relationship with elevated pulse pressure strenghtens over the adult lifespan, independently of overall blood pressure and heart rate variability^11^. These findings further support the hypothesis that that the white matter integrity mediates the relationship between pulse pressure and cognition, particularly processing speed and fluid intelligence.

Here, we tested whether the effects of pulse pressure on cognition are mediated by PSMD. We used data from a large-scale, lifespan population-based cohort of healthy adults. Our primary hypothesis was that the relationship between pulse pressure and processing speed is mediated by PSMD, even after accounting for other key indicators of vascular health like global blood pressure levels and heart rate variability. Secondary hypotheses included that the mediation effect (i) increased with age and (ii) have consequences for fluid intelligence too. To test these hypotheses, we used three latent vascular factors, predominantly expressing: pulse pressure, steady state blood pressure and heart rate variability^11^. This latent approach provides better estimates estimates of the underlying vascular constructs than individual measurements alone, offering a more robust test of our hypotheses^29–32^. We related these to white matter microstructure and cognition in a three-step process of: 1) developing linear and mediation models; 2) repeating the models over age sub-groups; 3) and expanding the models from processing speed to higher cognitive function.

## METHODS

### Participants

We studied participants in the population-based Cam-CAN cohort, which has deeply phentotyped data on approximately 700 adults, aged 18-88 years^33,34^. Figure 1 illustrates the analytical approach. The methods were conducted in accordance with guidelines approved by Cambridgeshire 2 (now East of England—Cambridge Central) Research Ethics Committee (reference: 10/H0308/50), who approved all experimental protocols. All participants gave full, informed, written consent. Participants were recruited from Cambridge City GP surgeriess, randomly to help maximise population representativeness of the cohort. The detailed recruitment pathway is outlined elsewhere^33^. Particpants were cognitively healthy and free from a history of dementia, referral for dementia assessment or memory complaints, with Mini-Mental State Examination >24/30^35^, and revised Addenbrokes Cognitive Examination score >82/100. Education was reported across four categories of: none, GCSE or O-Level, A-Level, Degree (College or University).

**Figure 1.**
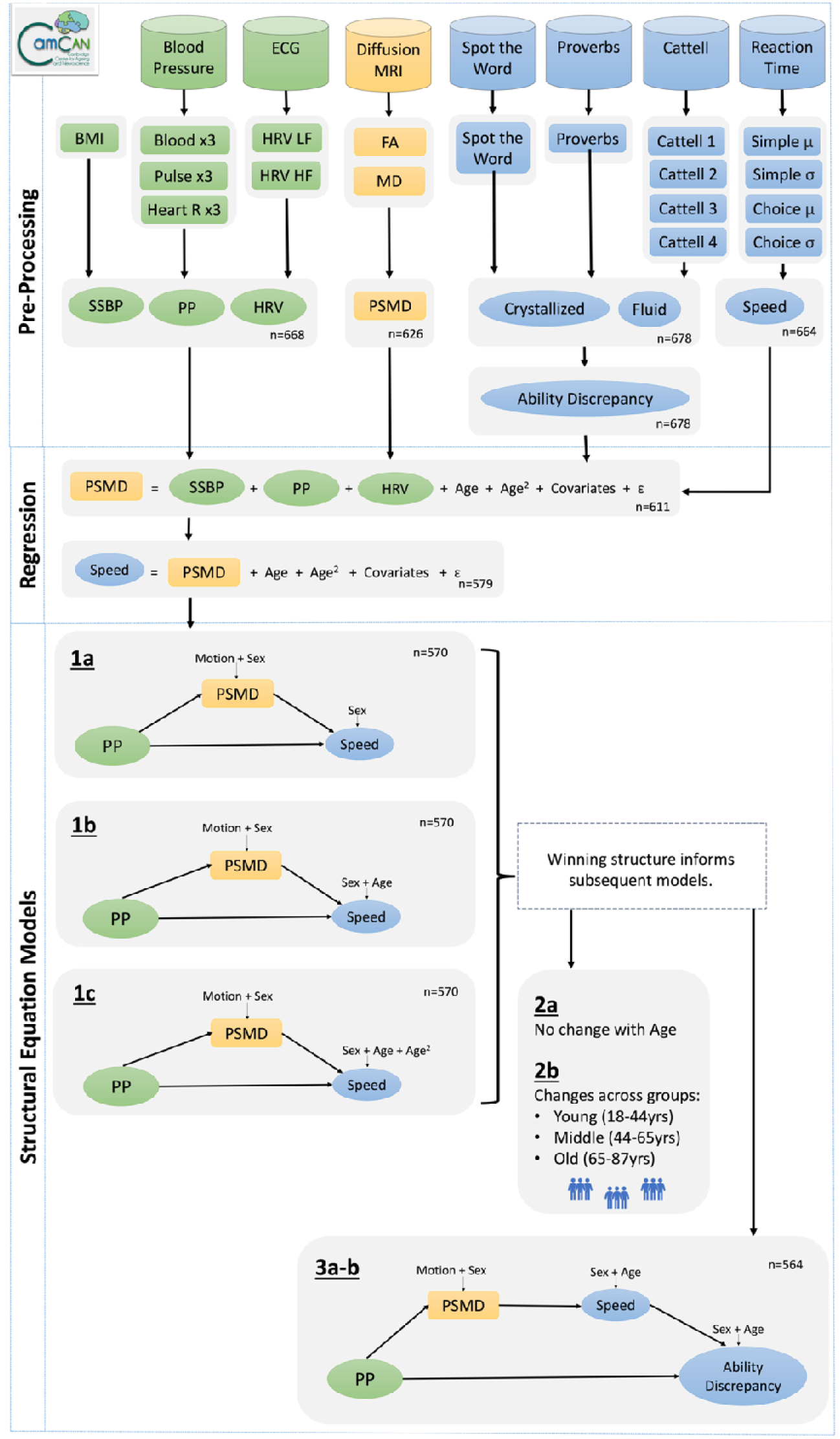
A schematic representation of the main stages in the data processing and analysis pipeline, to investigate whether PSMD, an indicator of white matter disruption, mediated the relationship between pulse pressure (systolic – diastolic) and cognitive decline, in the Cam-CAN dataset (n ≤ 708). For simplicity, the schematic does not show the quadratic latent vascular factors. Abbreviations: Blood, blood pressure (systolic + diastolic); BMI, body mass index; ECG, electrocardiogram; FA, fractional anisotropy, Heart R, heart rate; HRV HF, heart rate variability high frequency; HRV LF, heart rate variability low frequency; MD, mean diffusivity; PSMD, peak width of skeletonized mean diffusivity; PP, pulse pressure (systolic – diastolic); SSBP, steady state blood pressure. Symbols: µ, mean; σ, standard deviation

### Cardiovascular Measures and Latent Vascular Factors

Observations were recorded for body mass index, heart rate, heart rate variability at low and high frequencies, and systolic and diastolic blood pressures. Medication status (binary on/off) was reported for drugs with cardiovascular relevance, across four categories: [1] anti-hypertensives; [2] beta blockers; [3] other diuretics; [4] dyslipidemics. Full details of these observations are available elsewhere^19^.

The vascular observations were processed following our previous methodology^19^, as summarised in Figure 1. In brief, the three repeated observations of systolic and diastolic blood pressures were used to calculate three iterations of steady state blood pressure (systolic + diastolic), and pulse pressure (systolic – diastolic). These three observations were condensed into a single latent variable, as were three observations of mean heart rate (pulse rate). The resulting variables were modelled, alongside body mass index and heart rate variability at low and high frequencies. These six vascular observations formed three latent vascular factors using Exploratory Factor Analysis. Steady state blood pressure loaded strongly onto the first latent factor, with a small contribution from body mass index. Pulse pressure loaded strongly onto the second latent factor, with a small negative contribution from mean heart rate. The measures of heart rate variability in both high and low frequencies loaded similarly onto the third latent factor. Each latent factor is referred to subsequently by the variable name with most prominent loading. The pulse pressure factor was the focus of the present theory-driven analysis, but to explore whether pulse pressure acts independently to other vascular signals, the factor scores for all three latent vascular factors were extracted and input to the statistical models outlined below. Sensitivity analysis examined whether the results remained consistent when using observed measures of pulse pressure instead of latent vascular factors,

### Behavioural Tasks and Cognitive Measures

Assessments of crystallized and fluid intelligence were also outlined previously^19^. In brief, six observed measures were condensed into two latent variables, representing crystallized and fluid intelligence (n=678). The difference between these latent cognitive factors was calculated to give the “ability discrepancy” score^36^. The ability discrepancy was based on three assumptions: (1) fluid and crystallized intelligence measurents are age-invariant, (2) the two are highly correlated in youth, and (3) crystallized measures remains stable with age; as previously motivated^11^.

Processing speed was captured through response times in “simple” and “choice” tasks. In both tasks, the outcome measure was the time between presentation of a visual cue and pressing a button with a finger of the right hand. In the simple task, there was only one type of cue, to which participants responded with their index finger only. In the choice task, different cues indicated which of four fingers to use. Full details are in ^33^. The mean and standard deviation of response times were calculated across the 40-60 trials with correct responses. Both reaction time tasks (in milliseconds) were positively skewed, therefore were log-transformed to better align with Gaussian distributions. The sign of the transformed scores was then flipped, such that higher scores represented faster responses, consistent with previous approaches^37,38^. Finally, speed scores were standardised (Z-scored), and condensed into a single latent variable representing processing speed, using the “cfa” function in the lavaan package^39^. Missing data (simple task, N=1; choice task, N=1) were imputed using Full Information Maximum Likelihood, in cases where data were recoded for at least two observed variables, producing latent factor scores for n = 664. Factor score estimates for the latent variable were extracted for further analyses, below.

### Diffusion Tensor Imaging and White Matter Microstructure Measures

Pre-processing of the MRI data used the SPM12 software (Wellcome Department of Imaging Neuroscience; https://www.fil.ion.ucl.ac.uk/spm), release 4537, implemented in the Automatic Analysis pipeline, release 4.2^34,40^. In brief, 1mm isotropic T1- and T2-weighted images were bias-corrected for inhomogeneity of the magentic field, segmented, and warped to match a gray matter template created from the whole CamCAN sample using SPM’s DARTEL toolbox. This template was subsequently affine transformed to standard MNI space. Details of the MR sequences are available here: https://camcan-archive.mrc-cbu.cam.ac.uk/dataaccess/pdfs/CAMCAN700_MR_params.pdf, and an XML summary of AA preprocessing is available here: https://camcan-archive.mrc-cbu.cam.ac.uk/dataaccess/ImagingScripts/mri_aa_release004_roistreams_v1_tasklist.xml.

The 2mm isotropic diffusion weighted imaging (DWI) were processed with a common pipeline that has been described in detail elsewhere^41^. In brief, DWI data were pre-processed including correction for noise and Gibbs ringing using DIPY tools (https://dipy.org/), eddy current distortions, and head movement using eddy in FSL (https://fsl.fmrib.ox.ac.uk/fsl/fslwiki/). After these pre-processing steps, six datasets were excluded from further analysis: two due to corrupted DWI, resembling ‘salt and pepper’, and four due to excessive motion artefacts. Correction for B0 field inhomogeneities was not applied because reverse phase-encode direction data was not available. DTI fitting was performed by excluding the b=2000 s/mm^2^ data, using weighted linear least squares fitting in FSL, and mean diffusivity (MD) and fractional anisotropy (FA) maps were generated.

The FA and MD maps were used to calculate the global peak width of skeletonised mean diffusivity (PSMD), following previous work^12^, shown in Figure 2. In brief, all participant’s FA maps were skeletonized, using the standard Tract-Based Spatial Statistics procedure, in FSL^42^. This created a group-specific mean FA template and a white matter skeleton. Each participant’s FA map was normalised to the group-specific FA template and projected onto the skeleton template (Figure 2, top row). The transformation and projection parameters were then applied to the participant’s MD map using tbss_non_FA script, creating a corresponding skeleton of MD values in group-specific space (Figure 2, bottom row). To avoid contamination of the MD skeleton by partial volume effects, particularly from cerebrospinal fluid, the MD skeleton was masked using the group-specific mean FA skeleton, thresholded above an FA value of 0.3. This tends to restrict voxels to cortical regions and exclude voxels close to ventricles, as advised^12^. In the resulting masked MD skeleton, the value of each voxel represents an MD value. A greater spread of MD values indicates global white matter breakdown.

**Figure 2.**
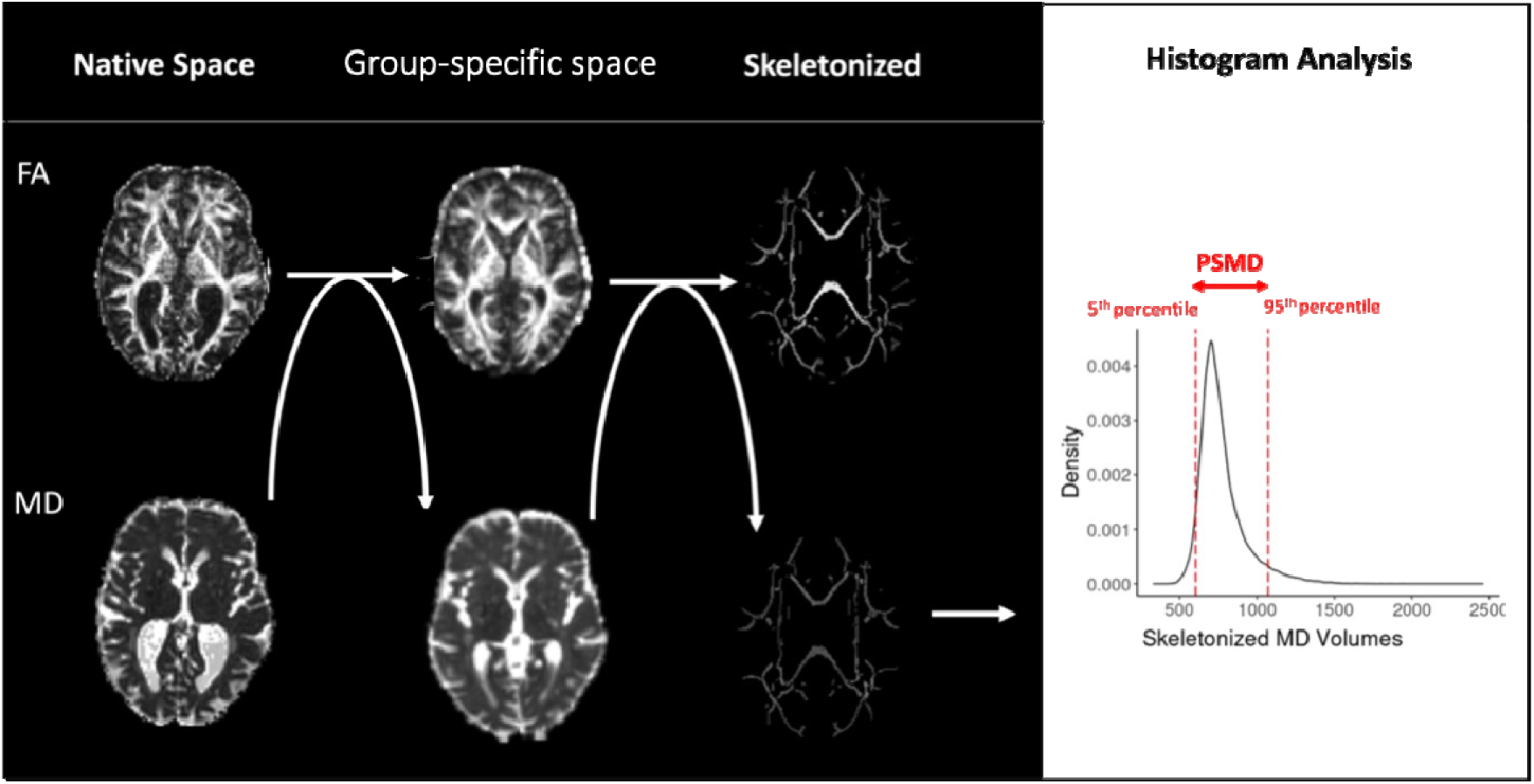
An illustration of the main steps in the procedure to calculate the peak width of skeletonised mean diffusivity (PSMD). The diffusion tensor imaging metrics of fractional anisotropy (FA) and mean diffusivity (MD) values (mm^2^/s) were normalised to MNI space. Normalised FA values were then projected onto a skeleton template. The skeletonization projection parameters were next applied to the normalised MD values. The skeletonized and masked MD values were input to histogram analysis (right). The difference between the 5^th^ and 95^th^ percentiles (red dashed lines) was calculated, to give PSMD. Higher PSMD indicates greater heterogeneity in white matter integrity, potentially indiciating diffuse white matter damage.

For each participant, the distribution of their MD values, across voxels within the masked skeleton, were plotted as a histogram (Figure 2). The difference between the 95th and 5th percentiles of the MD value was calculated (and divided by 1×10^6^ to convert to mm^2^/s, as in previous studies). A low PSMD value indicates greater uniformity in MD values throughout the white mater. A higher value indicates greater heterogeneity, assumed to reflect diffuse disruption across one or more white matter tracts. To understand how the calculated PSMD values corresponded to the input MD maps, we visualised the calculation process for three example participants (Figure 3).

**Figure 3.**
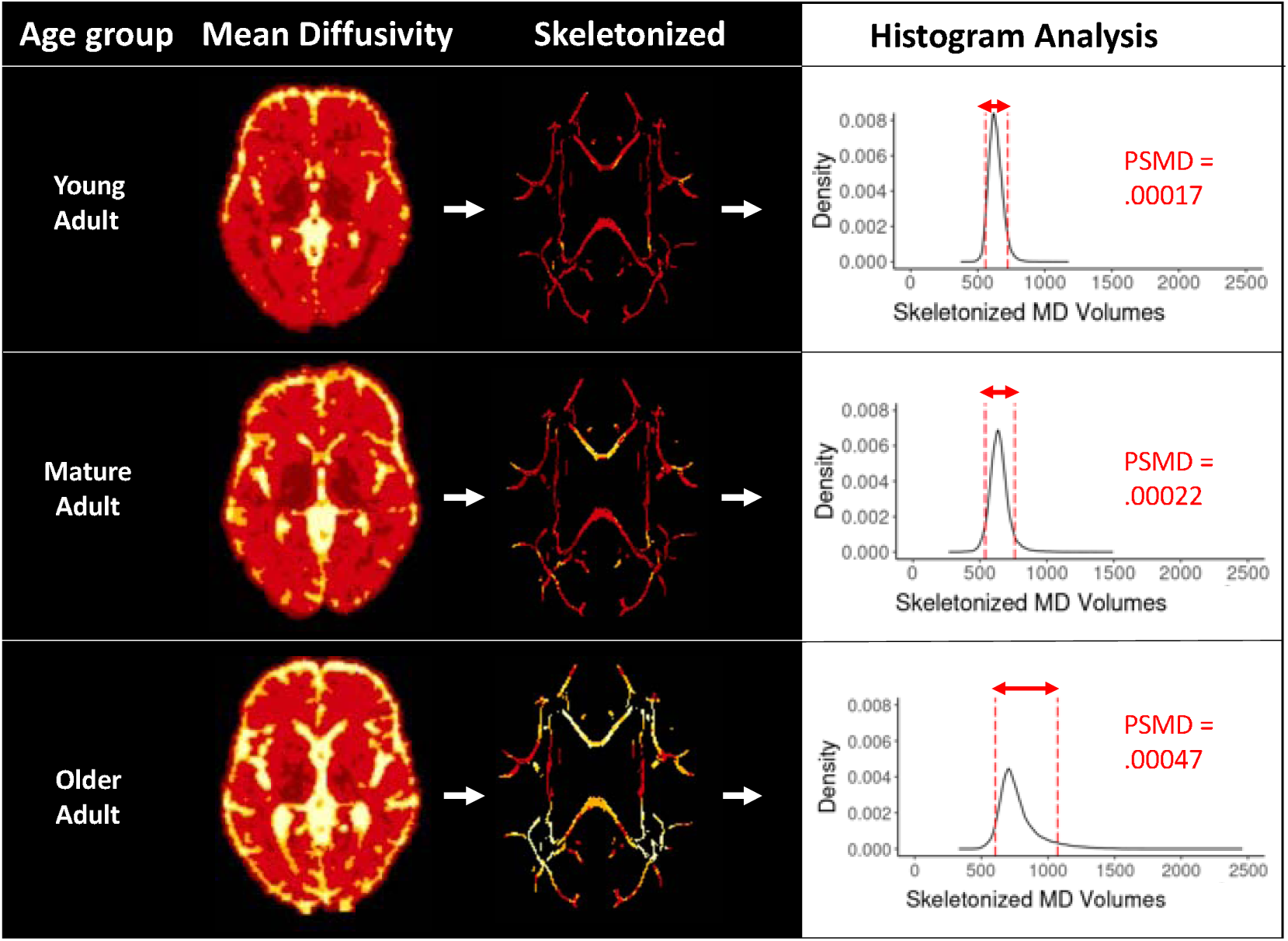
An illustration of the main steps in calculating of the peak width of skeletonised mean diffusivity (PSMD) for three example participants: a young adult in early twenties, a middle-aged adult in their late fifties, and an old adult in their late eighties. The mean diffusivity images (left), here normalised to standard space, are coloured red to yellow indicating low to high mean diffusivity values (mm^2^/s). High diffusivity is shown in yellow. In the skeletonized images (middle), a greater range of colours within an image suggests a greater range of mean diffusivity values. The distribution and density of mean diffusivity values were plotted on histograms (right). The difference between the 5^th^ and 95^th^ percentiles (red dashed lines) was used to calculate PSMD. Higher PSMD indicates non-uniform white matter integrity. Here, PSMD values were lowest in the youngest participant and highest in the oldest participant.

DWI is sensitive to head motion, which creates artefacts that reduce diffusion estimates and tend to increase with age. To account for this, stripe index was used to estimate head motion effects and included as a covariate in subsequent analyses^43^.

### Analytical Integration of Vascular, White Matter and Cognitive Measures

Pairwise relationships between vascular, cerebral and cognitive measures were examined using Pearson’s product-moment correlation coefficients. Since pulse pressure appeared to have a quadratic relationship with PSMD (Error! Reference source not found.), latent vascular factors were additionally considered in their quadratic forms in the subsequent regression and SEM models. All variables were standardised (mean=0, standard deviation=1) before input to regression and SEM models.

Outliers with undue influence motivated the use of robust linear regression, using the MASS package^44,45^. We performed a series of regression models from simple to complex, only retaining variables that benefited model fit. Model fit was investigated with the Akaike Information Criterion (AIC), Bayesian Information Criterion (BIC) and proportion of variance explained. Results were deemed significant if p<0.05. Predictor p-values were adjusted with Bonferroni corrections for the larger models with over twenty predictors.

Regression models established evidence for relationships between pulse pressure and PSMD, and between PSMD and processing speed (See Supplemental Section A), this motivated formally testing whether the effects of pulse pressure on PSMD drive individual differences in processing speed, i.e, whether an association between pulse pressure and processing speed is *mediated* by PSMD^46^. This was tested in a series of SEMs using the lavaan package^39^. Schematic representations of mediation models are shown in Figure 1. The base directly connects the predictor (green) on the left to the dependent variable (blue) on the right. The diagonal paths are referred to as “a” and “b”, which together form the indirect mediating pathway (“a x b”). The total modelled effect of the predictor onto the outcome is denoted as “c”, while after accounting for the mediating pathway, the remaining variance in the direct path is denoted as “c’”.

To calculate the proportion of the total effect that is explained by the indirect pathway, we divided the indirect path estimate by the total effect, i.e, (a x b) / (a x b + c’). This was complicated by instances where the direct and indirect effects counteracted one another – so-called “suppressor effects” – such that the total effect was less than the sum of the absolute effects. To overcome this, we used absolute values ^47^.

The SEMs included linear and quadratic forms of age and pulse pressure, for completeness. In this case, the total mediation effect was reported as the sum of the absolute linear and quadratic pulse pressure mediation pathways: (a_1_ x b) + (a_2_ x b).

Model structure was based on theory and results of regression models. Model fit was assessed for the initial model, using RMSEA and its confidence interval, CFI and SRMR. Good fit was defined as RMSA<0.05, CFI>0.97, and SRMR<0.05^48^. Statistical inferences on subsequent adaptations to model structure were made by comparing nested models via the likelihood ratio chi-squared difference test (p<0.05). Model paths were considered significant if the bootstrapped (n=5,000) confidence intervals did not cross zero^49^.

SEM 1a explored whether the relationship between pulse pressure and processing speed (direct path, c’) was mediated by PSMD (indirect path, a x b), above the covariates of sex and head motion (see Figure 1). Both linear and quadratic expressions of pulse pressure were used as mediators.

SEM1a was next expanded to account for linear age in SEM 1b. The need to include linear age was assessed by comparing SEM 1b to a version in which the path between Speed and age was constrained to be equal to zero. Age was taken forwards only if it improved model fit. This process was repeated for the inclusion of quadratic age in SEM 1c.

The winning model version from SEM 1b-c informed the specification of an additional checking step, SEM 1d. This model investigated whether the mediation pathway was specific to pulse pressure, over and above other vascular signals. SEM 1d included linear and quadratic steady state blood pressure and heart rate variability factors.

Next we used multigroup SEM (SEM 2a-b) to investigate whether the mediation effects (in the winning model from the SEM 1 models) varied with age. It is possible that the strength of the entire mediation pathway changed across the lifespan in this broad sample (18-87 years). In our previous study^19^, pulse pressure had unique effects on cognitive ability discrepancy (the difference between crystallised and fluid intelligence) only in older adults. Here, the cohort was categorized into three equally sized sub-groups (n=190) of young (18-44 years), middle (44-65 years) and old (65-87 years) adults. The structure of SEM 1a was repeated, now additionally using the Lavaan argument for multiple sub-groups. In SEM 2a, all model paths were constrained to be equal across age groups, whereas in SEM 2b, the ‘a_1_’ and ‘a_2_’ paths connecting pulse pressure linear and quadratic to PSMD, were allowed to vary across the three age groups. SEM 2a-b were then compared.

In the third set of models (SEM 3a-b), the mediation pathway was expanded to test whether the effects of pulse pressure on processing speed contribute to cognitive ability discrepancy,which is an approximation of longitudinal decline in fluid intelligence, as outlined previously ^11^. This tested whether individual differences in processing speed underpin “downstream” differences in fluid intelligence ^28^. The model expansion was also motivated by previous findings that the ability discrepancy was significantly predicted by pulse pressure interacting with age ^11^. By including ability discrepancy, SEM 3a became a *dual mediation* model (a_1_ x b_1_ x b_2_), see Figure 1. To check replication of Kievit et al.^37^, SEM 3b was also run on fluid intelligence alone, rather than ability discrepancy. In an attempt to move closer to understanding the causal sequence of events, the final model was compared to a version where the order of processing speed and fluid intelligence (or ability discrepancy) was reversed, i.e. pulse pressure to PSMD to fluid intelligence to processing speed (but covariates were again unchanged).

### Data Code and Availability

The raw data are available on request from https://camcan-archive.mrc-cbu.cam.ac.uk/dataaccess/. Statistical analyses were performed in R (version 4.0.2) and R-Studio ^50^. PSMD was calculated using the release 1.8.2 (https://github.com/miac-research/psmd/releases) ^12^. CSVs for the summary measures and R code for regression models and SEMS are available at: https://github.com/DebsKing/Pulse_pressure_impairs_cognition_via_white_matter_disruption.git.

## RESULTS

### Participants

Characteristics of the 708 participants in the Cam-CAN Phase 2 are outlined in Supplementary **Error! Reference source not found.**, suggesting the cohort is cognitively healthy and free from dementia.

### PSMD

PSMD values are illustrated in the supplementary material (Figure S1). PSMD and linear pulse pressure correlated strongly and positively (r = 0.43, p < 0.001), with a relationship that appeared to be non-linear (Figure S1). This motivated including quadratic vascular factors in analyses. Quadratic vascular effects are visualised in Figure S4.

**Figure 4.**
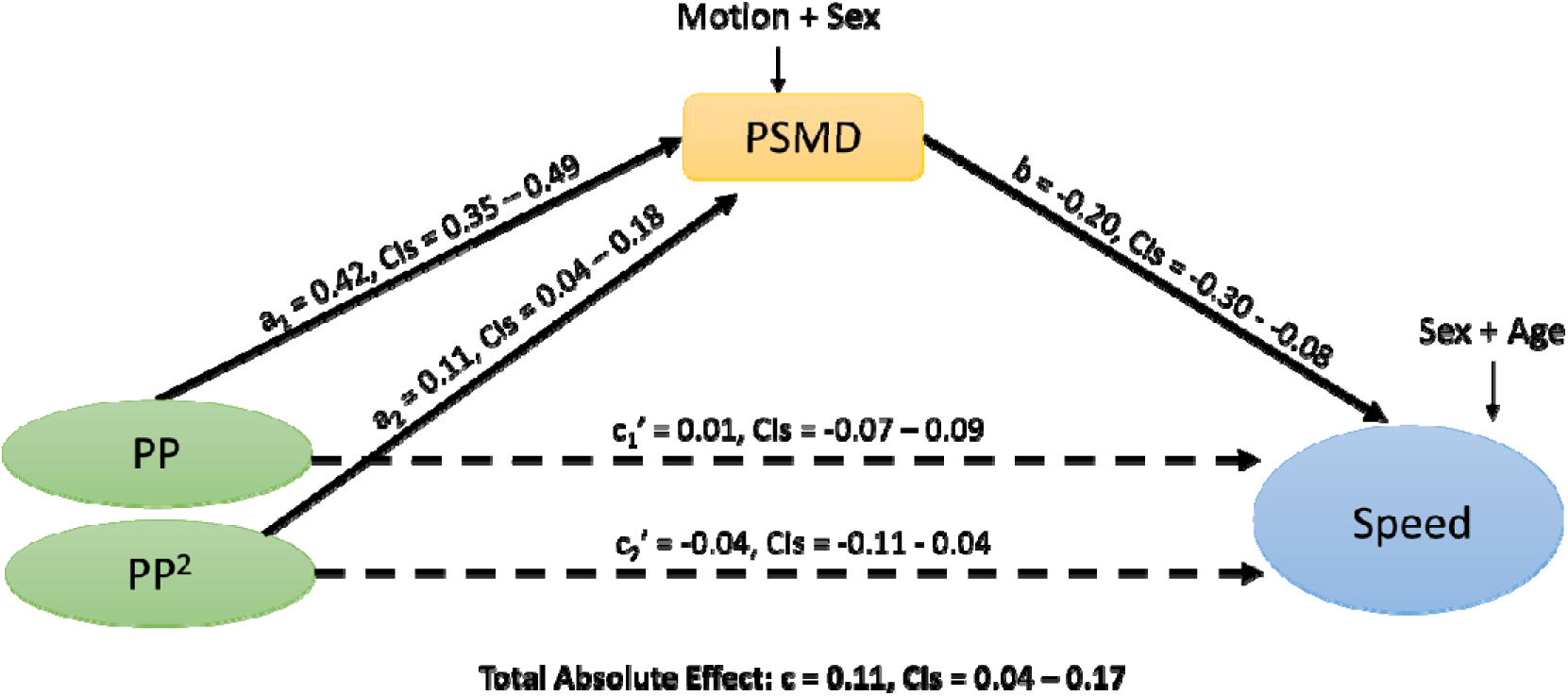
Structural Equation Model 1b (n=570) with standardized betas. Dashed lines represent insignificant results. Abbreviations: PP, pulse pressure; PSMD, peak width of skeletonized mean diffusivity, CIs = 95% bootstrapped confidence intervals.

### Structural Equation Models

The relationships between pulse pressure and PSMD, and between PSMD and processing speed, were significant in the linear regression models (Supplementary Section B). This motivated exploring whether the relationship between pulse pressure and processing speed is mediated by PSMD. We investigated this using a series of structural equation models, of increasing complexity (see Methods Section: Analytical Integration of Vascular, White Matter and Cognitive Measures).

SEM 1a modelled linear and quadratic pulse pressure contributions to processing speed, via PSMD with a good fit (Table S12 Table S12. Structural Equation Model 1a. Significant effects where confidence intervals do not cross zero are in bold.). In SEM 1a, the total absolute effect of linear and quadratic pulse pressure on processing speed was significant (c=0.41, CIs=[0.33–0.52]). This relationship was significantly mediated by PSMD (indirect absolute effect: ((a_1_ x b) + (a_2_ x b)) =0.29, CIs=[0.22–0.37]), accounting for 70% of the variance in the pulse pressure-speed relationship. This mediation was partial, meaning that the combined direct effects of linear and quadratic pulse pressure on speed remained significant (c_1_’ + c_2_’ =0.12, CIs=[0.04–0.25]).

SEM 1b expanded the model to remove linear effects of age on processing speed (Figure 4, Table S13). The total absolute effect of linear and quadratic pulse pressure on processing speed remained significant (c=0.14, CIs=[0.09–0.28]), as did the mediation effect ((a_1_ x b) + (a_2_ x b))=0.10, CIs=[0.04–0.17]), accounting for 72% of the variance. This mediation was partial, in that the combined direct effects of linear and quadratic pulse pressure on speed remained significant (c_1_’ + c_2_’ =0.04, CIs=[0.01–0.16]). To assess the validity of including Age, SEM 1b was compared to a model where the path to Age was constrained to be equal to zero; the unconstrained model fit best (Table S14), showing the importance of adjusting for age.

SEM 1c included quadratic effects of age on processing speed (Table S15). The total absolute effect of linear and quadratic pulse pressures on processing speed was still significant (c=0.14, CIs=[0.08–0.27]), as was the mediation effect ((a_1_ x b) + (a_2_ x b))=0.09, CIs=[0.03–0.16]), accounting for 68% of the variance in the pulse pressure-speed relationship. SEM 1c was compared to a model where the path to Age^2^ was constrained, and the constrained model fit best (Table S16), consistent with regression analysis findings where Model 2b (excluding Age^2^) fit the data better than Model 2c (including Age^2^). Therefore Age^2^ was not taken forwads into SEM 1d.

In a specificity analysis, SEM 1d investigated whether the mediation pathway was specific to pulse pressure, over and above other vascular signals. SEM 1d included the steady state blood pressure and HRV factors, in linear and quadratic forms (Figure S5, Table S17). The total absolute effect of all vascular factors on processing speed was significant (c=0.31, CIss=[0.24–0.62]). Pulse pressure to processing speed was significantly mediated by PSMD (indirect absolute effect: ((a_3_ x b) + (a_4_ x b))=0.08, CIs=[0.03–0.13]), accounting for 24% of the variance in the pulse pressure-speed relationship. HRV to processing speed was also significantly mediated by PSMD (indirect absolute effect: ((a_5_ x b) + (a_6_ x b))=0.07, CIs=[0.03–0.12]), accounting for 23% of the variance in the pulse pressure-speed relationship. The results observed in SEM 1a-1d remained consistent when replacing the latent vascular estimates with observed measures of pulse pressure and steady state blood pressure (Table S18-Table S23).

**Figure 5.**
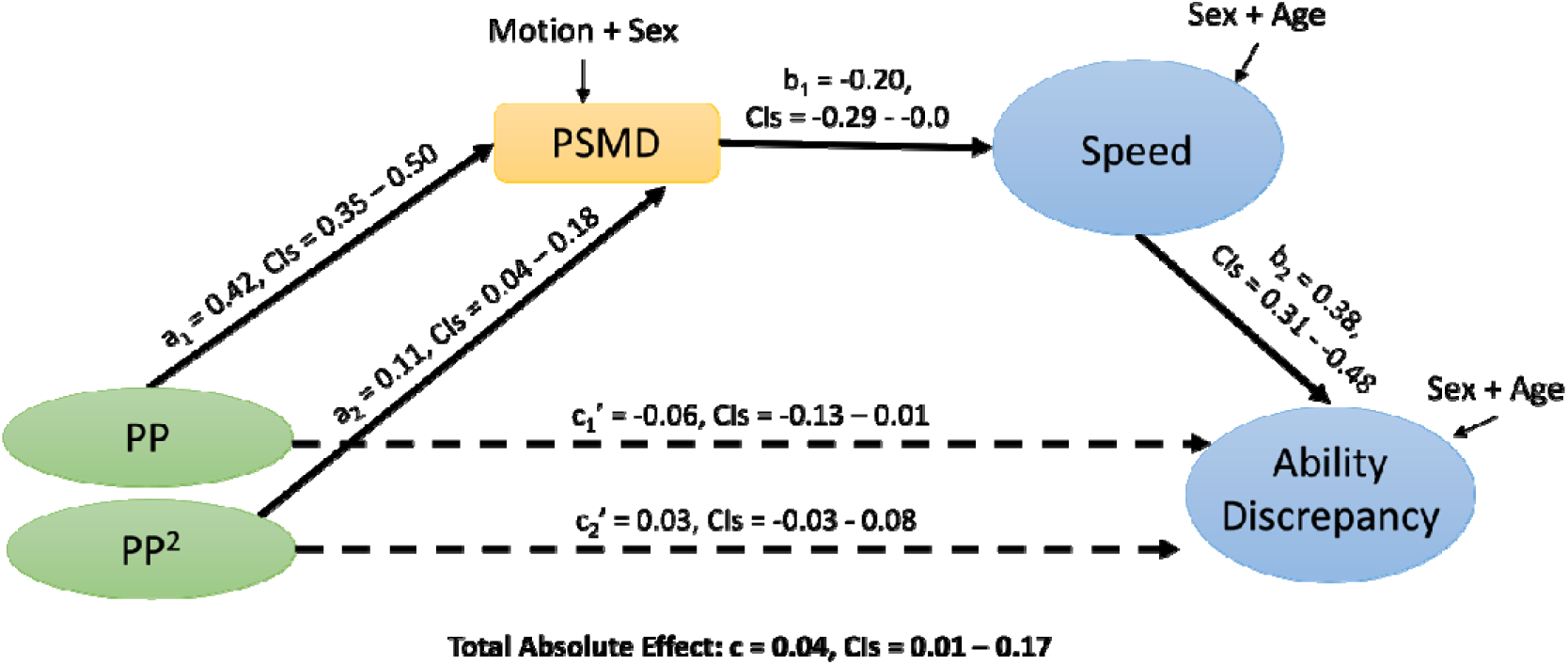
Structural Equation Model 3b (n=564). Shows standardized betas. Dashed lines represent insignificant results. Abbreviations: PP, pulse pressure; PSMD, peak width of skeletonized mean diffusivity.

It was possible that the strength of the entire mediation pathway changed with age, such that the effects of pulse pressure on white matter and cognition vary across the lifespan. To test this, we specified SEM 2a, where all models paths were constrained be equal across three age groups, of young (18-44 years), middle (44-65 years) and old (65-87 years) adults. This model was compared to SEM 2b, where the paths connecting pulse pressure to PSMD were allowed to vary across the three age groups. SEM 2a fit the data best (Table S24), indicating that the strength of the mediation pathway is stable across life adult lifespan.

The third set of models, motivated by the Wateshed model, tested whether the effects of processing speed had downstream effects on higher cognition. SEM 3a tested whether the effects of pulse pressure on PSMD related to the ability discrepancy score, a proxy for longitudinal decline in fluid intelligence. To test whether the ability discrepancy was required, SEM 3a was compared to a version where the ‘b_2_’ path from processing speed to the ability discrepancy was constrained to zero, and the full model fit best (Table S25), suggesting that speed did relate to ability discrepancy. In SEM 3a (Table S26), the total absolute effect of pulse pressure on the ability discrepancy was significant (c=0.03, CIs=[0.02–0.04]). However, evidence for dual-mediation of this relationship by PSMD and speed was borderline (indirect absolute effect: ((a_1_ x b) + (a_2_ x b))=0.01, CIs=[0.00–0.02]). As the Watershed model was originally established using measures of fluid intelligence rather than discrepancy scores^37^, we further examined whether the effects were significant when applied to fluid intelligence itself by testing SEM 3b.

Again, we first confirmed that the ‘b_2_’ path from processing speed to the fluid intelligence in SEM3b was needed, in that a model where it was constrained to zero was worse (Table S27). The total absolute effect of pulse pressure on fluid intelligence was significant (c=0.13, CIs=[0.06–0.21]) (Figure 5, Table S28), as was dual-mediation of this relationship by PSMD and speed (indirect absolute effect: ((a_1_ x b) + (a_2_ x b))=0.04, CIs=[0.01–0.07]), accounting for 31% of the variance in the pulse pressure-fluid intelligence relationship. The mediation was partial, in that the combined direct effects of linear and quadratic pulse pressure onto fluid intelligence remained significant (c_1_’ + c_2_’=0.09, CIs=[0.02–0.17]).

Finally, to get closer to causality, SEM 3b was compared to a model in which the direction of paths were reversed - such that fluid intelligence came before processing speed - using the “watershed” logic of ^28^. The expected order of variables fit best, irrespective of whether covariates are included or excluded (Table S29, Table S30).

## DISCUSSION

Using a multivariate approach we confirmed the hypothesis that the effects of pulse pressure on cognition are at least partly explained by the integrity of white matter microstructure. There were three key findings. Firstly, white matter microstructure significantly and substantially mediated the effect of pulse pressure on processing speed. Secondly, the strength of this mediation was stable across the adult lifespan. Thirdly, the effects of pulse pressure on procressing speed had downstream consequences for fluid intelligence.

These findings suggest that pulse pressure is a potential therapeutic target for reduced risk of cognitive decline and dementia^51^. We suggest that there is now sufficient evidence to motivate longitudinal and interventional studies to test whether better managing pulse pressure helps maintain white matter microstructure and preserve cognitive abilities in later life.

### PSMD and processing speed: A Sensitive Marker of Neurocognitive Ageing

The PSMD values in Cam-CAN were similar to those in comparable population-based cohorts^13,15^, and showed a similar profile of continuous increase with age, accelerating after 60 years^13,15^. Here, the spread of PSMD values also increased substantially with age. This reinforces previous suggestions that PSMD may reflect individual differences in early brain ageing.

Here, we replicated previous studies showed that increased PSMD is associated with reduced processing speed^12,21–24^. Moreover, the association remained after adjusting for linear and quadratice effects of age, suggesting that white matter integrity is similarly important for processing speed across the adult lifespan. Processing speed has special status in studies of cognitive ageing because it is thought to be the foundation for age-related differences in higher cognitive abilities^25–27^. Our findings reinforce evidence that PSMD is a sensitive marker of neurocognitive ageing, with additional potential relevance for dementia.

### Pulse Pressure and White Matter Microstructure

Pulse pressure correlated strongly and positively with PSMD, with a curvilinear relationship. The curve was relatively flat for low levels of pulse pressure, where incremental increases in pressure corresponded to only marginal increases in PSMD. The curve steepened almost exponentially at higher pressures. This could suggest that, while small fluctuations of normal levels in pulse pressure can be managed – for example through vessel elasticity and the windkessel effect – pressures beyond the limits of compensatory mechanisms penetrate the microcirculation, and drive cerebral damage with consequences for white matter integrity. Exponentially increasing damage could result from positive feed-back loops driven by high pulse pressure, such as high pressure causing vessel calcification and remodelling, which in turn raises the pressure further^1,51^.

In regression models, PSMD was uniquely predicted by quadratic pulse pressure, over and above other vascular factors. A relationship between pulse pressure and white matter is consistent with previous studies on vascular factors and white-matter hyperintensities^7,9,52^. Pulse pressure may impair white matter microstructure by damaging and remodelling the microvasculature, leading to hypoperfusion and in turn diffuse axonal loss, demyelination and inflammation^1^.

Independently to pulse pressure, the steady state blood pressure factor also predicted PSMD, although not as strongly. It highlights the multivariate nature and complexity of vascular ageing as previously argued^53^. Pulse pressure and steady state blood pressure predicted PSMD, independent of medications, supporting the idea that pulse pressure affects white matter beyond anti-hypertensive medications^8^. Here, while the linear model fit is best without medications, there were significant interactions. Pulse pressure interacted with beta-blockers and diuretics, while steady state blood pressure interacted with antihypertensives. While some of these could be false positives, given the number of terms (tests) in these models, they should be explored in future work aimed at understanding the underlying mechanisms and developing interventions for cerebrovascular ageing.

### Relating all three: Pulse Pressure to White Matter to Cognition

The effects of pulse pressure on PSMD, and of PSMD on processing speed, were examined simultaneously in mediation models. In the first model, PSMD accounted for 70% of the direct relationship between pulse pressure and processing speed. This is in line with previous work associating pulse pressure with white matter microstructure^7–10^, and white matter with cognitive performance^12,16,21,22^;^12,21–24^. The mediation effect remained significant and substantial when covarying age, and the strength of the mediation was stable across groups of young, middle and old aged participants. The importance of this result is best considered in the context of the cohort in which it was studied: The biggest risk factor for increasing pulse pressure is age^54,55^, and here, pulse pressure was studied in an adult cohort with a broad age range (18-88 years). For the mediation effect to be significant over and above age, in this cohort, suggests that there will be an even stronger effect when it is studied in a large sample with a smaller age range, such as in a group of middle-aged individuals in an interventional study.

A further analysis confirmed that pulse pressure affects white matter and cognition independently, over and above other vascular factors. However, there was additional significant and substantial mediation of HRV to processing speed. Low HRV has been linked to declines in processing speed in prior studies^56–58^. This further highlights the multifactorial nature of vascular ageing on brain health and cognitive function. Future work should explore whether pulse pressure and HRV contribute to loss of white matter integrity through similar or different mechanisms.

The model was next expanded to include the ability discrepancy, following the logic of the watershed hypothesis, namely that age-related declines in processing speed underpin downstream differences in higher cognition^28^. Mediation of pulse pressure on the ability discrepancy was borderline significant. Given that the watershed hypothesis was originally established using measures of fluid intelligence^28^, without adjusting for crystallised intelligence (as in the ability discrepancy score), we repeated this dual-mediation model with fluid intelligence as the outcome variable instead. Mediation of pulse pressure onto fluid intelligence, through PSMD and processing speed, was significant, even above age. The mediation pathway accounted for 31% of the total variance in fluid intelligence that was related to pulse pressure. Furthermore, reversing the paths (from fluid intelligence to pulse pressure) resulted in worse model fit. These results together support for the watershed hypothesis, and further highlight the importance of pulse pressure in cognitive ageing processes.

### Avenues to Treat Pulse Pressure

We speculate that better awareness and control of pulse pressure would slow the development and progression of white matter damage. Evidence from the regression models here suggests that beta-blockers and diuretics interact with pulse pressure, changing its effect on white matter microstructure. Further work is needed to explore the implications of these findings and to inform the development of new treatments. Interventions that are currently being tested include: reducing arterial siffness with aerobic exercise ^59^; lowering systolic blood pressure with anti-hypertensives, to improve cardiovascular and cognitive health, in the SPRINT-MIND trial^60,61^ and several multidomain interventions ^62^, including the FINGER trial, which aims to lower systolic blood pressure to benefit cognition in older adults ^63^.

### Strengths and Limitations

Cardiovascular, cerebral and cognitive ageing have often been studied in isolation. A key strength of this study is that we use an integrated, multivariate approach. To do this, we prioritised analysing high-quality data. The Cam-CAN cohort offers an unusual combination of multiple vascular and brain measures with high quality observations across multiple cognitive domains. This makes it an excellent “test bed” to develop new hypotheses about the multivariate effects of vascular ageing. Developing ideas in this way is a necessary precursor to rigorous testing in complex and resource-demanding longitudinal and intervention studies. However, it should be kept in mind that inferences drawn from our current findings could be limited by the cross-sectional design of currently available Cam-CAN data, in which age effects are counfounded by the effets of generational changes across people born in different years. When there is no temporal information, it is possible that the hypothesized direction - of ‘x’ causes ‘y’ via the mediator - is in fact reversed ^64^. Ideally, reverse causation would be theoretically implausible for any model developed in cross-sectional data. However, in our models, it is possible that instead of declines in cardiovascular health impairing cognitive abilities like processing speed, higher cognitive abilities in fact allow individuals to make lifestyle choices that improve their cardiovascular health. It also possible that a unidirectional relationship in either direction is overly simplistic. For example, regarding blood pressure, it was thought that hypertension caused secondary endothelial damage. However, there is some evidence that endothelial dysfunction happens first, and further evidence that both converge into a reinforcing and bidirectional relationship^65,66^. A similarly complex relationship may exist for pulse pressure and cognitive abilities, which would be difficult to untangle in cross-sectional data.

To try to address the possibility of reverse causation, we compared model fit with alternative orders of variables. The expected order – namely, pulse pressure to PSMD, to processing speed – fit the data best. However, this comparison is not conclusive about the true causal direction, and future work should extend this with longitudinal and interventional study designs. It is also possible that age drives changes in pulse pressure, which in turn drive changes in white matter and cognition. In this case, age could be modelled on the left of the vascular-brain-cognition triangle. Investigating this in future work will require longitudinal cohorts.

The latent vascular factor which we refer to as “pulse pressure” here was created using factor analysis, as in our previous study^11^, where it received small but possibly important contributions from heart rate variance. Importantly, the sensitivity analysis revealed that the results remained consistent when observed measures of pulse pressure were used instead of the latent vascular measures. While pulse pressure may only be a proxy for arterial stiffness, our approach of using a latent factor of pulse pressure may provide a more accurate representation of arterial stiffness. There is evidence that more direct measures, such as pulse wave velocity, explain greater variance in white matter hyperintensities^67^. Currently, the Heart and Brain study is collecting multiple measures of pulse pressure, alongside ultrasound metrics of vessel stiffness, white matter lesions and cognitive abilities^68^. These detailed measures of vascular pathology may contribute to better understanding the observed effects on brain and cognitive ageing. Thus future work should establish whether measuring only systolic and diastolic pressures is sufficient, or if a more comprehensive consideration of multiple vascular factors and modelling of their multifactorial nature is required. This will be an important step in translating the findings into clinical practice, where systolic and diastolic pressures (and heart rate) are more routinely measured.

The calculation of global PSMD, as used here, sacrifices detail on regional specificity. This may be important in light of evidence that some brain regions are more vulnerable to pulsatile forces. Future work could investigate tract-specific estimates of PSMD or other derived diffusion metrics, such as microstructural complexity^43^, and more conventional white-matter hyper-intensities that are routine in clinical MRI^69,70^.

### Conclusions

This study shows that white matter microstructure impairment mediates the effect of pulse pressure onto cognition. Better managing pulse pressure may help to preserve cognitive abilities throughout life.

## SUPPLEMENTAL MATRIAL

### Supplemental Section A: Methods

In total, ten regression models tested two sets of nested hypotheses. The first set of models sought to understand the relationship between PSMD and pulse pressure in the presence of other variables (see equations below). Model 1a examined which latent vascular factors make unique contributions to PSMD, when accounting for sex and head motion. Model 1b investigated whether any relationships between latent vascular factors and PSMD held above linear age, and Model 1c investigated whether any relationships held above linear and quadratic age. Since latent vascular factors were highly correlated with age, any loss of significance in Models 1b and 1c could be due to age sharing variance with these factors. Models 1a-c were compared, and if age terms improved fit, they were taken into Models 1d-e. Model 1d included medications relevant to cardiovascular health, as chosen and reported previously^19^. Medication status was treated as a series of categorical variables. Model 1e included interactions between vascular factors and sex, because of evidence for sex-related differences in the relationship between pulse pressure and white matter microstructure^8^. If model comparisons showed that the inclusion of medications and sex, and interactions between vascular factors, did not improve overall fit in Models 1d-e, these would not be taken forward to the SEMs, below. The multiple linear regression models are outlined in Supplemental Section A.

The multiple linear regression models are outlined below using Wilkinson’s notation ^71^. In this model syntax, latent vascular factors expressing steady state blood pressure, pulse pressure and heart rate variability are abbreviated to SSBP, PP and HRV, respectively; “β” are the parameter estimates (coefficients); “L” is the vector of residual errors; and interactions are indicated by “:”.

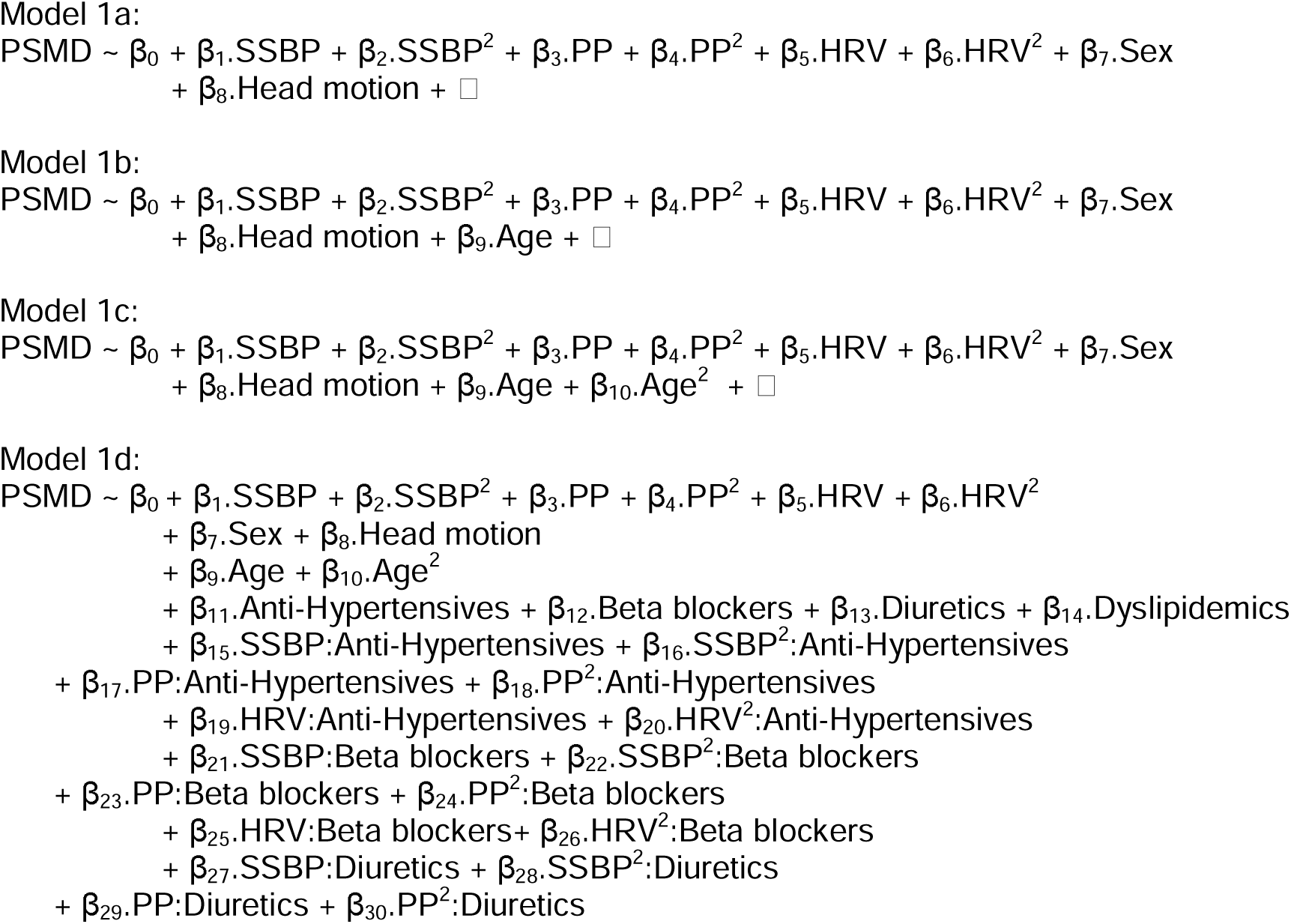
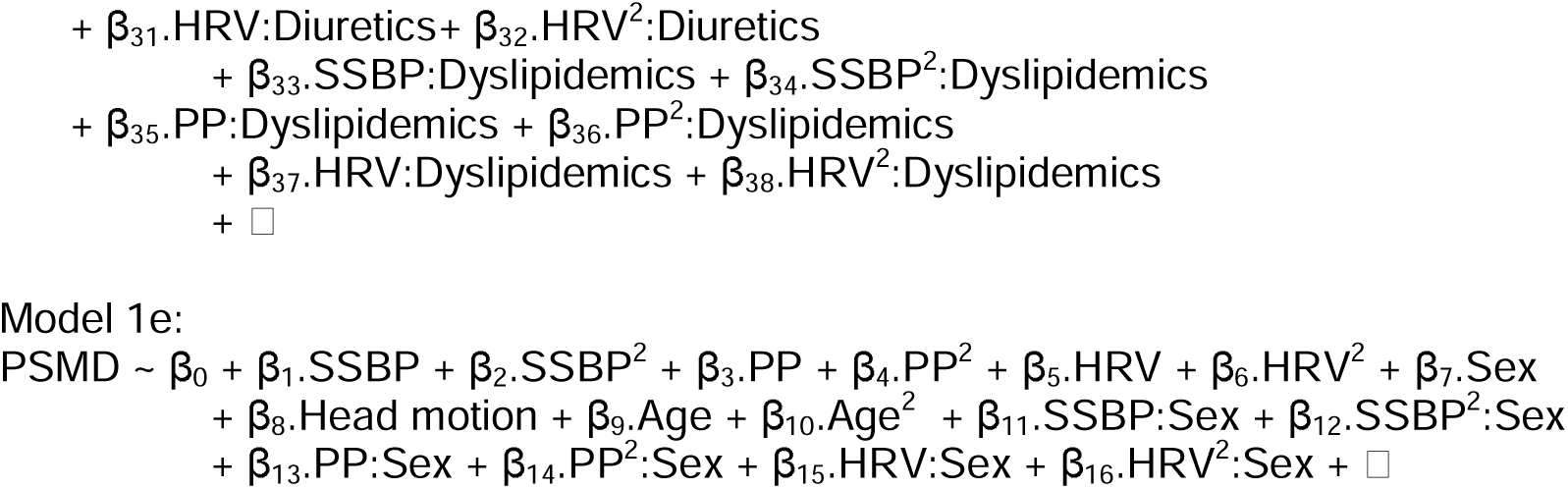

Regression models 2a-d attempted to replicate previous findings that PSMD related to processing speed ^12,21–24^. Model 2a accounted for sex and head motion. Model 2b additionally tested whether the effect of PSMD on speed held when accounting for linear age. Model 2c additionally accounted for quadratic age. If model comparisons showed additional terms did not improve model fit, these terms would not be taken into subsequent SEMs, below.

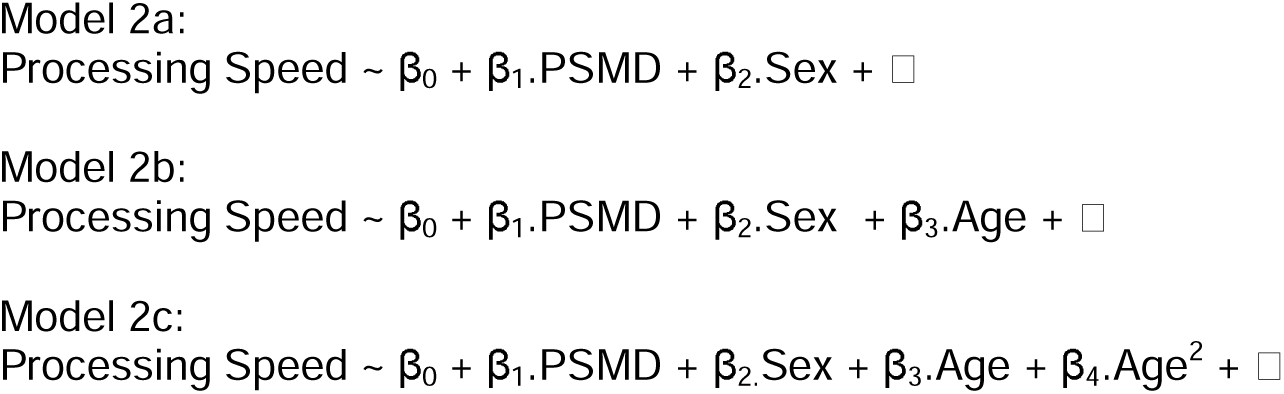

### Supplemental Section B: Results

Vascular observations were condensed into three latent vascular factors, using exploratory factor analysis, as in our previous study (King et al., 2023). The latent factors predominantly expressed steady state blood pressure, pulse pressure and heart rate variability. All latent vascular factors correlated significantly with age (Figure S1 **Error! Reference source not found.**).

**Figure S1.**
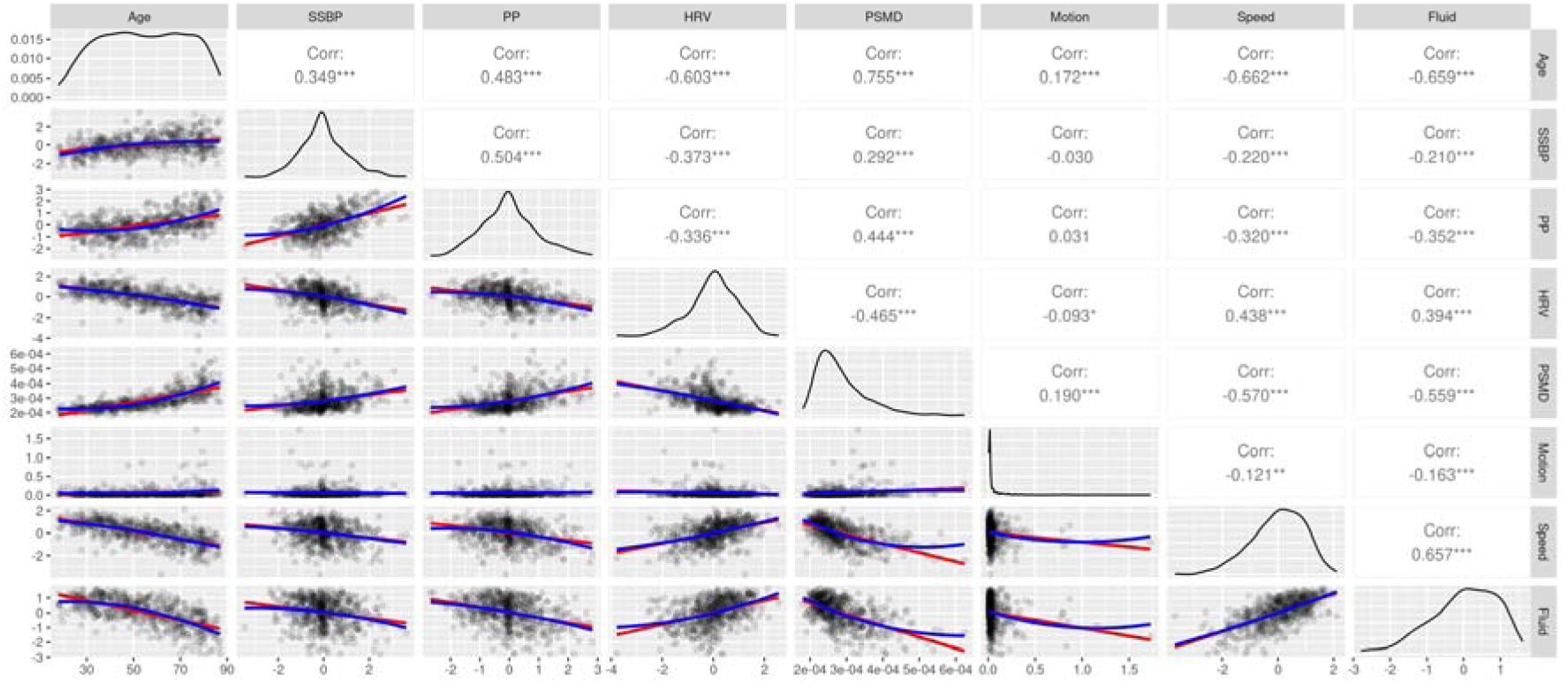
Scatter plots (lower left), distributions (leading diagonal) and Pearson correlations (upper right) for age, latent vascular factors, PSMD, head motion, processing speed and fluid intelligence (n=564). Scatter plots show linear (red) and quadratic (blue) associations and data intensity (greyscale). Stars indicate increasing significance on the correlations: ***, p<0.001; **, p<0.01; *, p<0.05.Abbreviations: SS BP, steady state blood pressure latent factor; Corr, correlation coefficient; HRV, heart rate variability latent factor; PSMD, peak width of skeletonised mean diffusivity; PP, pulse pressure latent factor.

The four RT observations (Figure S2) were condensed into a latent variable representing processing speed (Figure S3 **Error! Reference source not found.**). Processing speed and fluid intelligence both showed strong associations with age, and correlated significantly with the three latent vascular factors, with substantial effect sizes (Figure S1 **Error! Reference source not found.**).

**Figure S2.**
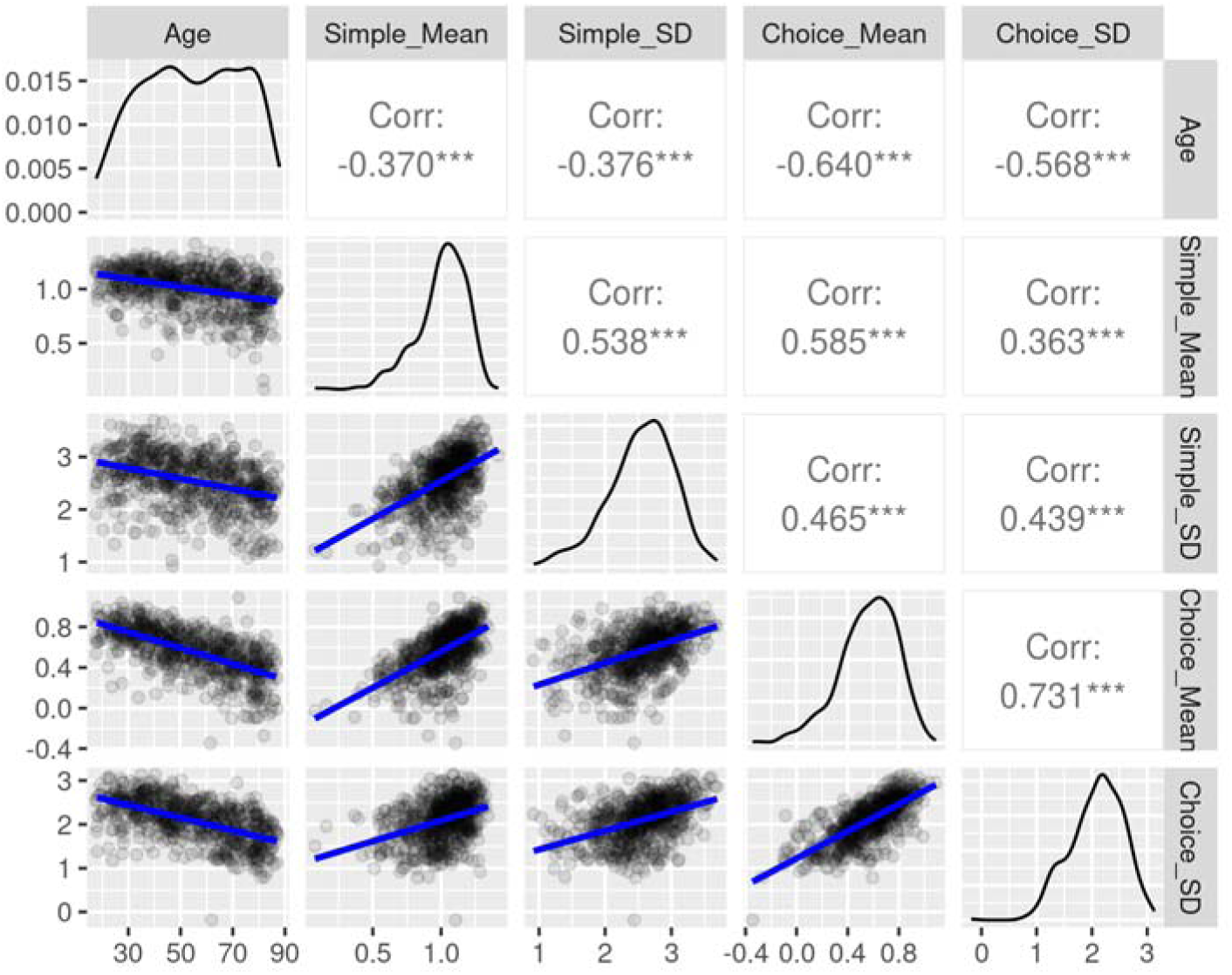
Scatter plots (lower left), distributions (leading diagonal) and Pearson correlations (upper right) for age and cognitive observed variables. Scatter plots show linear associations (blue) and data intensity (greyscale). Stars indicate increasing significance on the correlations: ***, p<0.001; **, p<0.01; *, p<0.05. Abbreviations: Choice_M, choice task mean; Choice_SD, choice task standard deviation; Corr, correlation coefficient; Simple_M, simple task mean; Simple_SD, simple task standard deviation.

**Figure S3.**
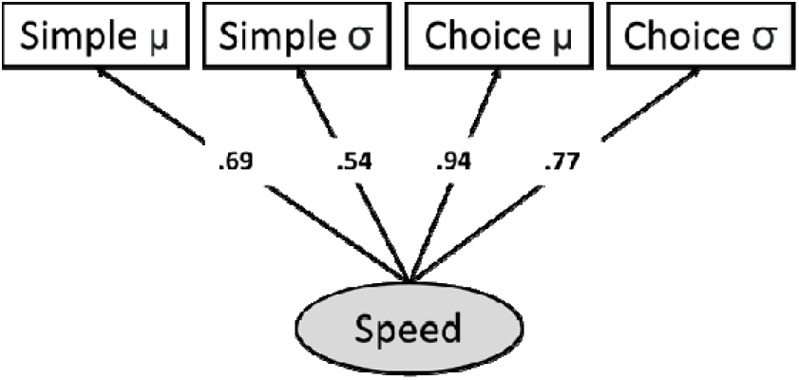
The one-factor Confirmatory Factor Analysis model of processing speed (n=664).

### Peak Width of Skeletonized Mean Diffusivity

PSMD was calculated for 620 participants. The values of PSMD are illustrated for three example participants in Figure 3. PSMD increased strongly with age (r = 0.75, p <0.001), with an uptick after 60 years of age (Figure S1). Head motion also increased with age (r = 0.16, p <0.001), and PSMD correlated moderately with head motion (r = 0.17, p <0.001). However, the correlation between PSMD and age remained strong after accounting for head motion (r = 0.75, p<0.001). Head motion was a covariate on PSMD in all subsequent analyses.

Correlations for complete case data across multimodal measures (n=564) are shown in Figure S1 **Error! Reference source not found.**.

### Multiple Linear Regressions

In the regression models predicting PSMD, Model 1a (Table S2 **Error! Reference source not found.**) showed a significant positive prediction from both linear pulse pressure (β = 0.25, p <0.001), and quadratic pulse pressure (β = 0.12, p <0.001). This was consistent with a relationship between pulse pressure and PSMD that was stronger at higher values of pulse pressure. There was a significant negative relationship with linear HRV (β = −0.33, p<0.001). PSMD also showed significant positive relationships with sex (β = 0.21, p<0.001) and head motion (β = 0.13, p<0.001).

The model was next expanded in three stages, first to include linear Age in Model 1b (Table S3). This improved model fit (Table S4), therefore linear Age was taken forwards into Model 1c, which additionally included quadratic Age (Table S5). This significantly improved model fit further, therefore linear and quadratic Age were taken into subsequent models. In Model 1d (Table S6), there were significant interactions between pulse pressure and both betablockers and diuretics, and between steady state blood pressure and anti-hypertensives. However, there were no improvements across fit indices when including medications in Model 1d, or interactions with sex in Model 1e (Table S7). Overall, Model 1c fit best.

In Model 1c (Table S5), PSMD was significantly and positively predicted by quadratic pulse pressure, over and above other vascular factors and linear and quadratic effects of Age (β = 0.05, p = 0.02). Head motion and sex also continued to show age-independent effects. Given these significant results, head motion, sex and both linear and quadratic effects of pulse pressure were taken into the pre-planned SEMs, below. There was also a significant effect of linear steady-state blood pressure (β = 0.05, p = 0.04); this was explored further in the post-hoc SEM, which included all latent vascular factors.

Taken together, the results of the regression Models 1a-e motivated including linear and quadratic expansions of pulse pressure in the subsequent SEM analyses. Additionally, since both linear and quadratic effects of Age explained significant variance in PSMD, above pulse pressure (in Model 1c), both age terms were also incorporated into the SEM analyses.

In the regression models predicting Processing Speed, Model 2a (Table S8) showed a significant negative relationship with PSMD (β = −0.62, p <0.001), and a positive relationship with sex (β = 0.29, p <0.001). These effects held above linear Age in Model 2b (Table S9), which improved model fit (Table S10), Model 2c with quadratic age fit worse (Table S11 **Error! Reference source not found.**). Overall, Model 2b fit best. Importantly, the effect of PSMD on Processing Speed remained significant over and above linear (Model 2b) and quadratic (Model 2c) effects of Age.

**Figure S4.**
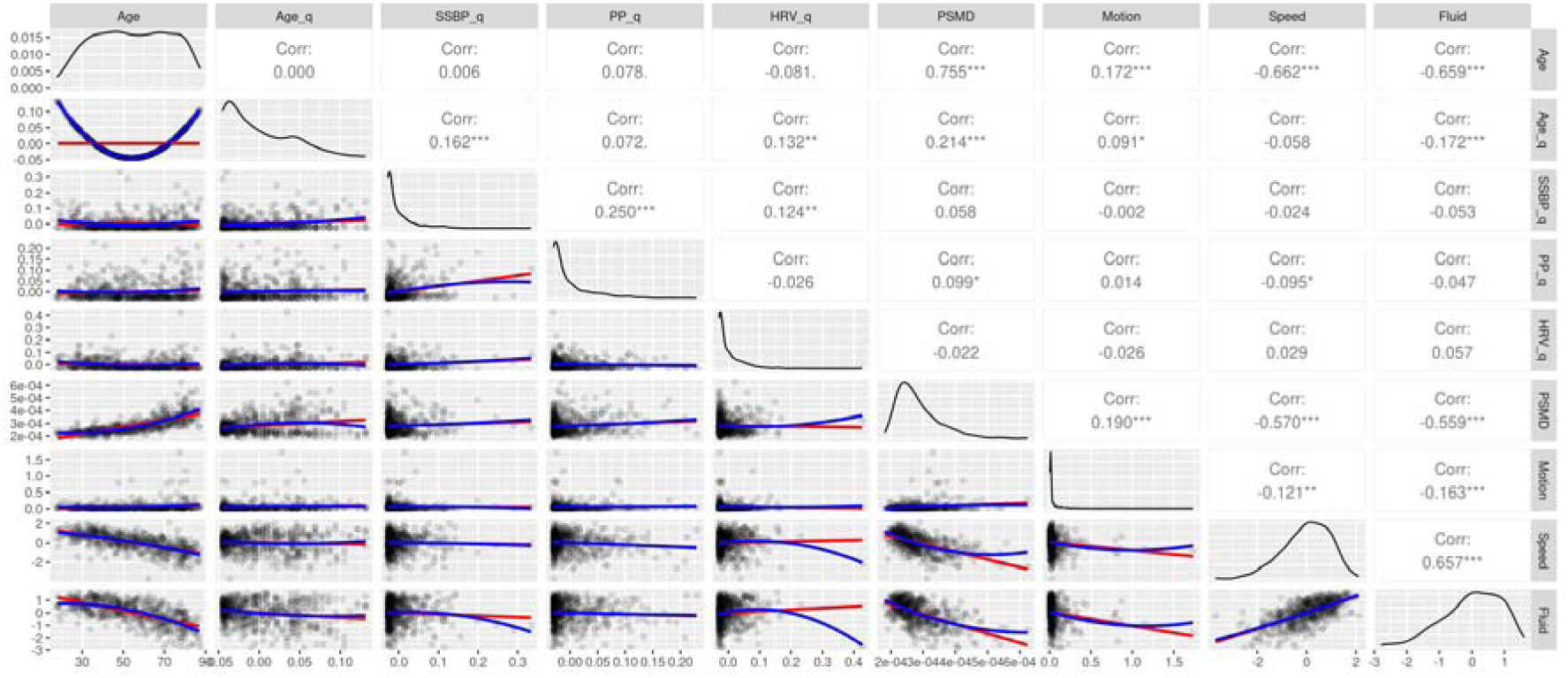
Scatter plots (lower left), distributions (leading diagonal) and Pearson correlations (upper right) for age, quadratic age, quadratic latent vascular factors, PSMD, head motion, processing speed and fluid intelligence (n=564). Scatter plots show linear (red) and quadratic (blue) associations and data intensity (greyscale). Stars indicate increasing significance on the correlations: ***, p<0.001; **, p<0.01; *, p<0.05. Variables named “xx_q” are in quadratic form.

**Figure S5.**
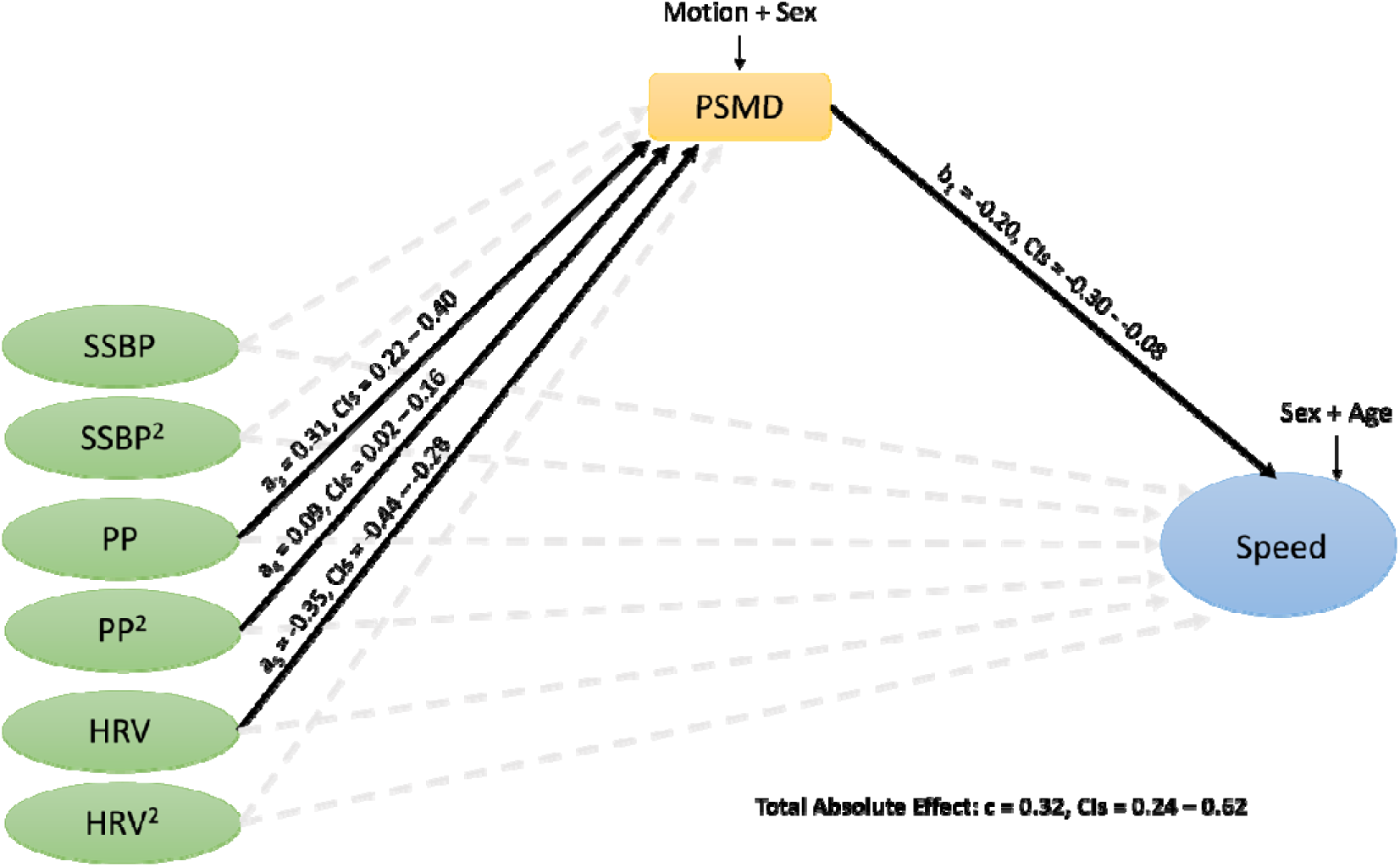
Structural Equation Models 1d (n=570). Standardized betas shown for significant paths only, for clarity. Dashed lines represent insignificant results. Abbreviations: HRV, heart rate variability; SSBP, steady state blood pressure; PP, pulse pressure; PSMD, peak width of skeletonized mean diffusivity.

### Supplemental Tables

**Table S1.** Demographic information for 708 participants, by equally split age groups. One decimal place is reported where data are continuous

|  | Complete Data (n) |  |  | Missing (%) |  |  | Range / Number (Male) |  |  | Mean |  |  | SD |  |  |
| --- | --- | --- | --- | --- | --- | --- | --- | --- | --- | --- | --- | --- | --- | --- | --- |
|  | Young | Middle | Old | Young | Middle | Old | Young | Middle | Old | Young | Middle | Old | Young | Middle | Old |
| Age (years) | 236 | 236 | 236 | 0 | 0 | 0 | 18-45 | 45-66 | 66-88 | 33.1 | 54.9 | 75.9 | 7.2 | 6.4 | 5.8 |
| Sex (Male) | 236 | 236 | 236 | 0 | 0 | 0 | 115 | 110 | 124 | - | - | - | - | - | - |
| MMSE | 235 | 236 | 235 | 0.4 | 0 | 0.4 | 25-30 | 26-30 | 25-30 | 30 | 29.1 | 29 | 1 | 1.1 | 1 |
| Education | 235 | 236 | 235 | 0.4 | 0 | 0.4 | - | - | - | - | - | - | - | - | - |
| (percentage of total with complete data, by category) |  |  |  |  |  |  |  |  |  |  |  |  |  |  |  |
| No qualifications tried (< 16) | 0.4 | 3.4 | 15.7 | - | - | - | - | - | - | - | - | - | - | - | - |
| GCSEs / O-levels (age 16) | 11.5 | 14 | 15.3 | - | - | - | - | - | - | - | - | - | - | - | - |
| A-levels (age 18) | 14.5 | 20.8 | 23.4 | - | - | - | - | - | - | - | - | - | - | - | - |
| Degree (over 18) | 73.6 | 61.9 | 45.5 | - | - | - | - | - | - | - | - | - | - | - | - |

Abbreviations: GCSE, The General Certificate of Secondary Education; MMSE, Mini-Mental State Examination; SD, standard deviation.

**Table S2.** Results of Model 1a (n = 611, DoF = 602 residual standard error = 0.60) with dependent variable PSMD. Significant effects (p<0.05) are shown in bold.

| Predictors | Standard $\beta$ | Standard Error | Confidence Intervals | P |
| --- | --- | --- | --- | --- |
| SSBP | 0.01 | 0.03 | -0.06 – 0.07 | 0.84 |
| SSBP<sup>2</sup> | 0.00 | 0.03 | -0.05 – 0.06 | 0.95 |
| PP | 0.25 | 0.03 | 0.19 – 0.31 | <0.001 |
| PP<sup>2</sup> | 0.12 | 0.03 | 0.07 – 0.18 | <0.001 |
| HRV | -0.33 | 0.03 | -0.38 – -0.27 | <0.001 |
| HRV<sup>2</sup> | 0.03 | 0.03 | -0.03 – 0.08 | 0.31 |
| Head motion | 0.13 | 0.03 | 0.07 – 0.18 | <0.001 |
| Sex | 0.21 | 0.06 | 0.10 – 0.32 | <0.001 |

**Table S3.** Results of Regression Model 1b (n = 611, DoF = 601, residual standard error = 0.49) with dependent variable PSMD. Significant effects (p<0.05) are shown in bold.

| Predictors | Standard $\beta$ | Standard Error | Confidence Intervals | p |
| --- | --- | --- | --- | --- |
| SSBP | 0.00 | 0.02 | -0.05 – 0.05 | 0.92 |
| SSBP<sup>2</sup> | 0.04 | 0.02 | 0.00 – 0.09 | 0.05 |
| PP | 0.07 | 0.03 | 0.02 – 0.12 | 0.01 |
| PP<sup>2</sup> | 0.07 | 0.02 | 0.03 – 0.11 | <0.01 |
| HRV | -0.01 | 0.03 | -0.06 – 0.04 | 0.64 |
| HRV<sup>2</sup> | 0.04 | 0.02 | -0.00 – 0.08 | 0.07 |
| Head motion | 0.06 | 0.02 | 0.02 – 0.10 | <0.01 |
| Sex | 0.26 | 0.04 | 0.18 – 0.34 | <0.001 |
| Age | 0.61 | 0.03 | 0.56 – 0.67 | <0.001 |

**Table S4.** Results of comparisons on Models 1a to 1d, using AIC and BIC, and the sum of squares derived from ANOVA comparisons.

|  | Difference in AIC | Difference in BIC | Difference in Sum Of Squares |
| --- | --- | --- | --- |
| Model 1a vs 1b | 298.21 | 293.80 | 159.00 |
| Model 1b vs 1c | 72.70 | 68.29 | 28.84 |
| Model 1c vs 1d | 1.69 | -121.94 | 19.98 |
| Model 1c vs 1e | -8.86 | -35.35 | 1.14 |

**Table S5.** Results of Regression Model 1c (n = 611, DoF = 600, residual standard error = 0.46) with dependent variable PSMD. Significant effects (p<0.05) are shown in bold.

| Predictors | Standard $\beta$ | Standard Error | Confidence Intervals | p |
| --- | --- | --- | --- | --- |
| SSBP | 0.05 | 0.02 | 0.00 – 0.10 | 0.04 |
| SSBP<sup>2</sup> | 0.04 | 0.02 | -0.00 – 0.08 | 0.07 |
| PP | 0.00 | 0.03 | -0.05 – 0.05 | 0.87 |
| PP<sup>2</sup> | 0.05 | 0.02 | 0.01 – 0.09 | 0.02 |
| HRV | 0.02 | 0.02 | -0.03 – 0.06 | 0.54 |
| HRV<sup>2</sup> | 0.02 | 0.02 | -0.02 – 0.06 | 0.33 |
| Head motion | 0.04 | 0.02 | 0.00 – 0.08 | <0.05 |
| Sex | 0.28 | 0.04 | 0.20 – 0.36 | <0.001 |
| Age | 0.68 | 0.03 | 0.63 – 0.73 | <0.001 |
| Age<sup>2</sup> | 0.19 | 0.02 | 0.15 – 0.23 | <0.001 |

**Table S6.**
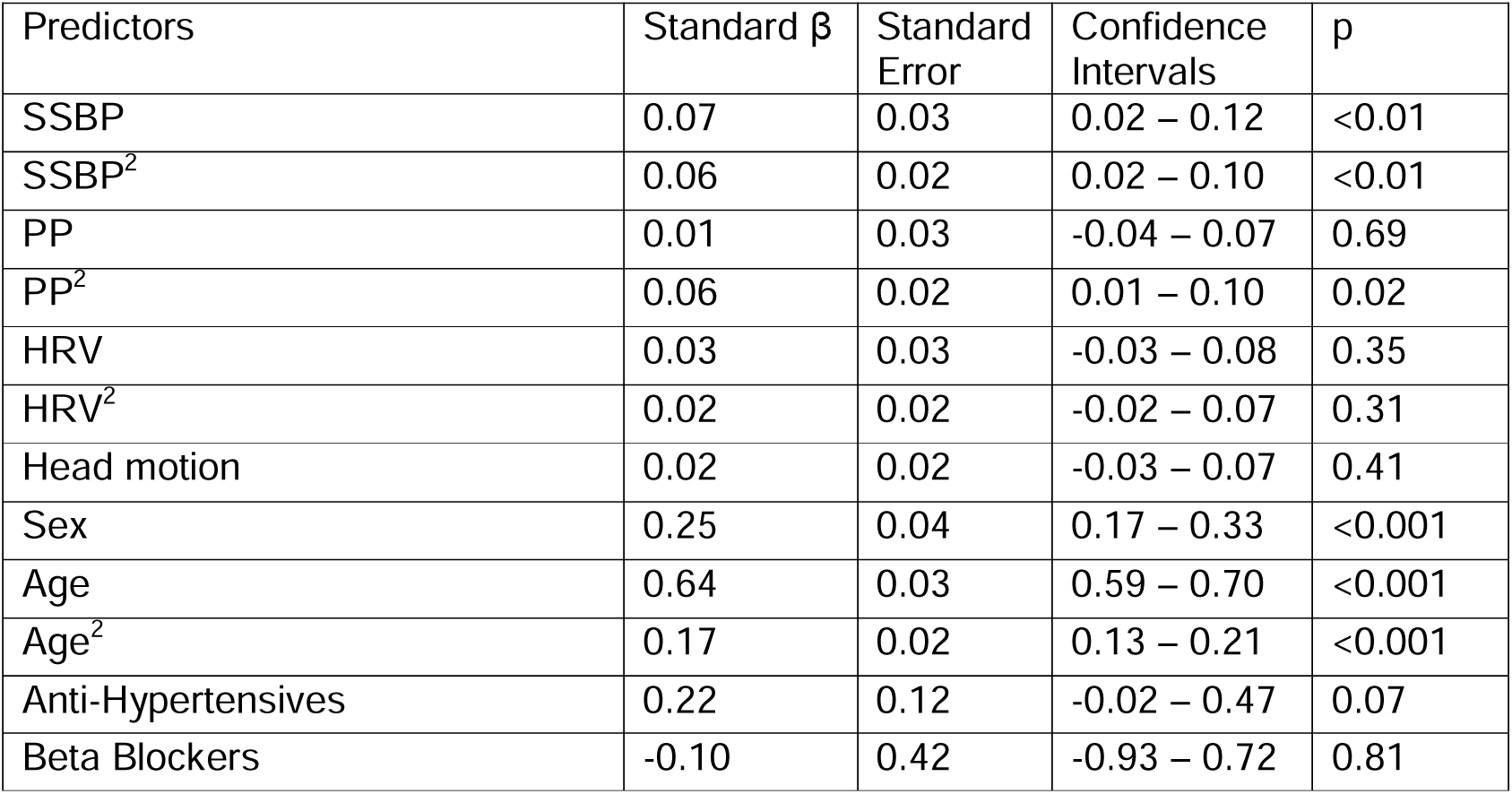
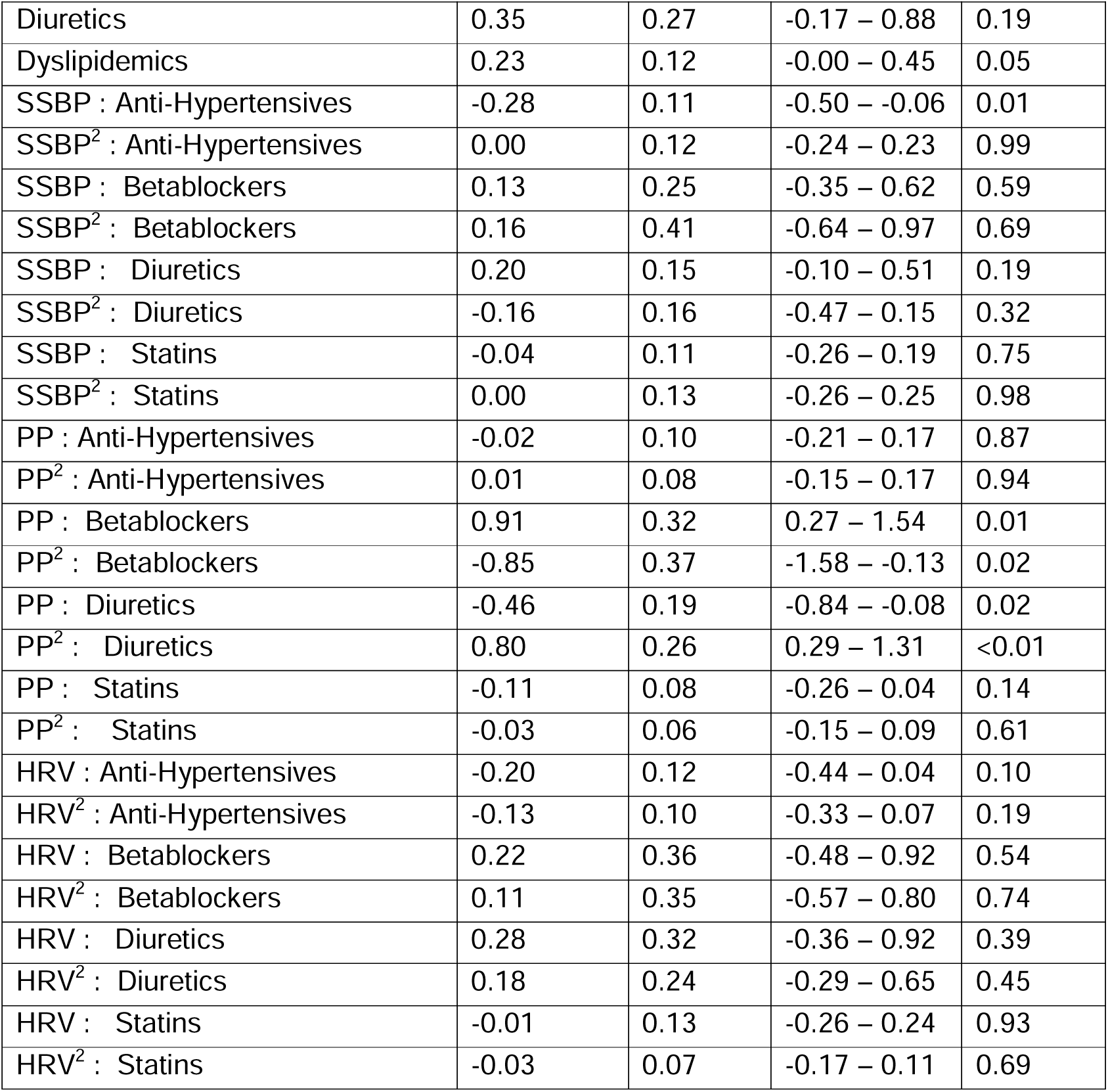
Results of Regression Model 1d (n = 611, DoF = 572, residual standard error = 0.42) with dependent variable PSMD. Significant effects (p<0.05) are shown in bold. The adjusted p-value with Bonferroni corrections for this non-winning model, containing 38 orthogonal tests, would be p<0.0013.

| Predictors | Standard $\beta$ | Standard Error | Confidence Intervals | p |
| --- | --- | --- | --- | --- |
| SSBP | 0.07 | 0.03 | 0.02 – 0.12 | <0.01 |
| SSBP<sup>2</sup> | 0.06 | 0.02 | 0.02 – 0.10 | <0.01 |
| PP | 0.01 | 0.03 | -0.04 – 0.07 | 0.69 |
| PP<sup>2</sup> | 0.06 | 0.02 | 0.01 – 0.10 | 0.02 |
| HRV | 0.03 | 0.03 | -0.03 – 0.08 | 0.35 |
| HRV<sup>2</sup> | 0.02 | 0.02 | -0.02 – 0.07 | 0.31 |
| Head motion | 0.02 | 0.02 | -0.03 – 0.07 | 0.41 |
| Sex | 0.25 | 0.04 | 0.17 – 0.33 | <0.001 |
| Age | 0.64 | 0.03 | 0.59 – 0.70 | <0.001 |
| Age<sup>2</sup> | 0.17 | 0.02 | 0.13 – 0.21 | <0.001 |
| Anti-Hypertensives | 0.22 | 0.12 | -0.02 – 0.47 | 0.07 |
| Beta Blockers | -0.10 | 0.42 | -0.93 – 0.72 | 0.81 |
| Diuretics | 0.35 | 0.27 | -0.17 – 0.88 | 0.19 |
| Dyslipidemics | 0.23 | 0.12 | -0.00 – 0.45 | 0.05 |
| SSBP : Anti-Hypertensives | -0.28 | 0.11 | -0.50 – -0.06 | 0.01 |
| SSBP<sup>2</sup> : Anti-Hypertensives | 0.00 | 0.12 | -0.24 – 0.23 | 0.99 |
| SSBP : Betablockers | 0.13 | 0.25 | -0.35 – 0.62 | 0.59 |
| SSBP<sup>2</sup> : Betablockers | 0.16 | 0.41 | -0.64 – 0.97 | 0.69 |
| SSBP : Diuretics | 0.20 | 0.15 | -0.10 – 0.51 | 0.19 |
| SSBP<sup>2</sup> : Diuretics | -0.16 | 0.16 | -0.47 – 0.15 | 0.32 |
| SSBP : Statins | -0.04 | 0.11 | -0.26 – 0.19 | 0.75 |
| SSBP<sup>2</sup> : Statins | 0.00 | 0.13 | -0.26 – 0.25 | 0.98 |
| PP : Anti-Hypertensives | -0.02 | 0.10 | -0.21 – 0.17 | 0.87 |
| PP<sup>2</sup> : Anti-Hypertensives | 0.01 | 0.08 | -0.15 – 0.17 | 0.94 |
| PP : Betablockers | 0.91 | 0.32 | 0.27 – 1.54 | 0.01 |
| PP<sup>2</sup> : Betablockers | -0.85 | 0.37 | -1.58 – -0.13 | 0.02 |
| PP : Diuretics | -0.46 | 0.19 | -0.84 – -0.08 | 0.02 |
| PP<sup>2</sup> : Diuretics | 0.80 | 0.26 | 0.29 – 1.31 | <0.01 |
| PP : Statins | -0.11 | 0.08 | -0.26 – 0.04 | 0.14 |
| PP<sup>2</sup> : Statins | -0.03 | 0.06 | -0.15 – 0.09 | 0.61 |
| HRV : Anti-Hypertensives | -0.20 | 0.12 | -0.44 – 0.04 | 0.10 |
| HRV<sup>2</sup> : Anti-Hypertensives | -0.13 | 0.10 | -0.33 – 0.07 | 0.19 |
| HRV : Betablockers | 0.22 | 0.36 | -0.48 – 0.92 | 0.54 |
| HRV<sup>2</sup> : Betablockers | 0.11 | 0.35 | -0.57 – 0.80 | 0.74 |
| HRV : Diuretics | 0.28 | 0.32 | -0.36 – 0.92 | 0.39 |
| HRV<sup>2</sup> : Diuretics | 0.18 | 0.24 | -0.29 – 0.65 | 0.45 |
| HRV : Statins | -0.01 | 0.13 | -0.26 – 0.24 | 0.93 |
| HRV<sup>2</sup> : Statins | -0.03 | 0.07 | -0.17 – 0.11 | 0.69 |

**Table S7.**
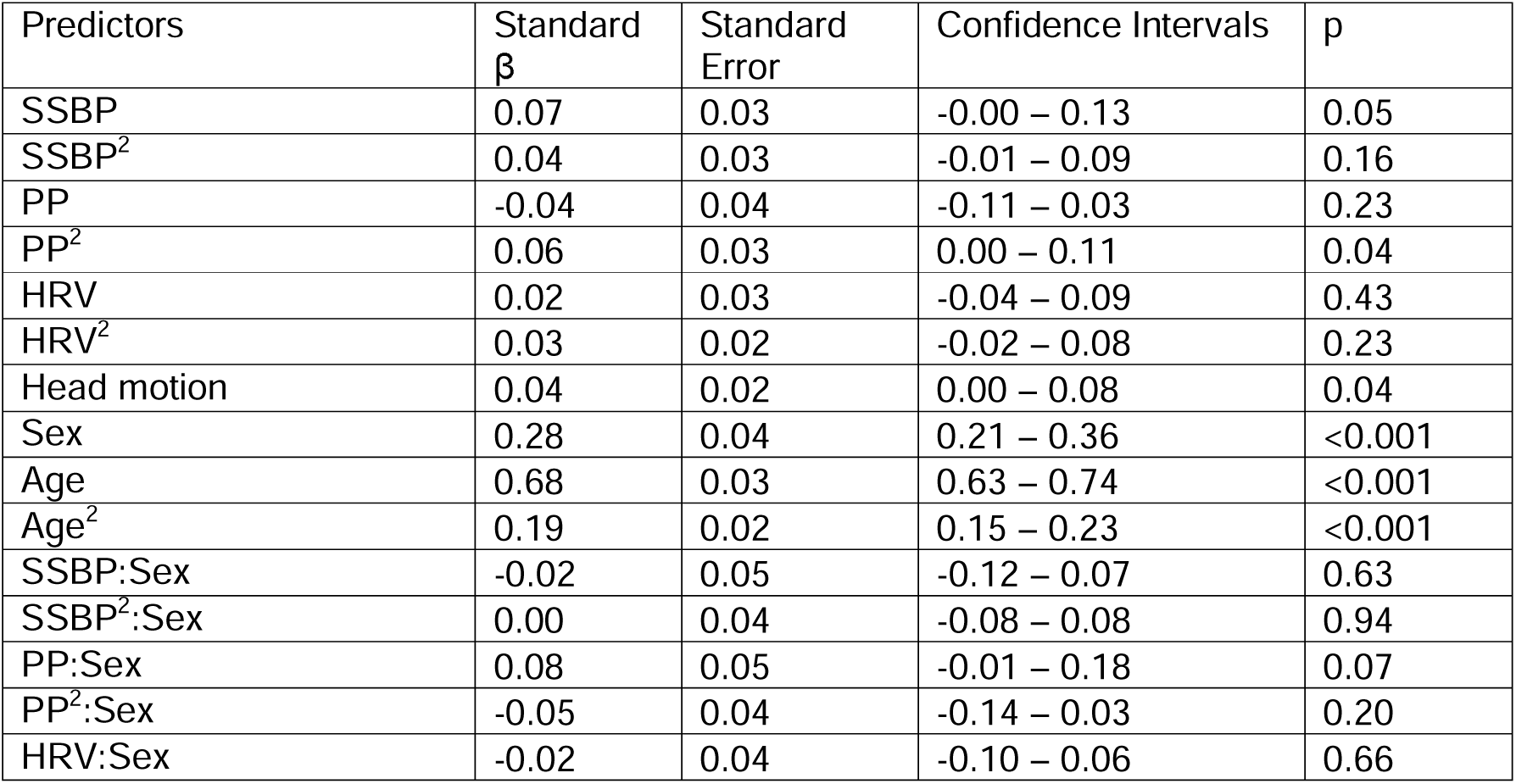
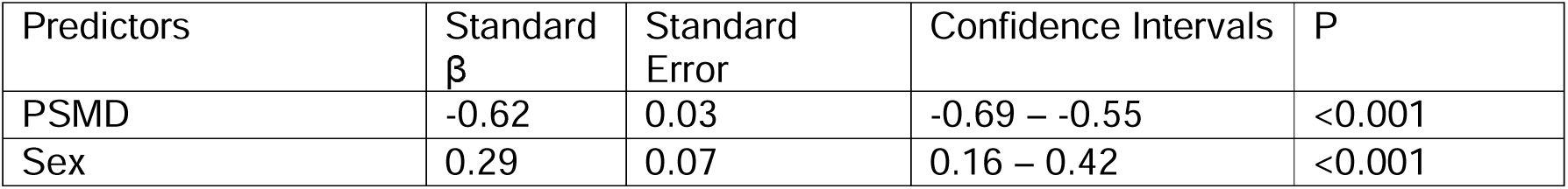
Results of Regression Model 1e (n = 611, Do F = 594, residual standard error = 0.44) with dependent variable Processing Speed. Significant effects (p<0.05) are shown in bold.

**Table S8.**
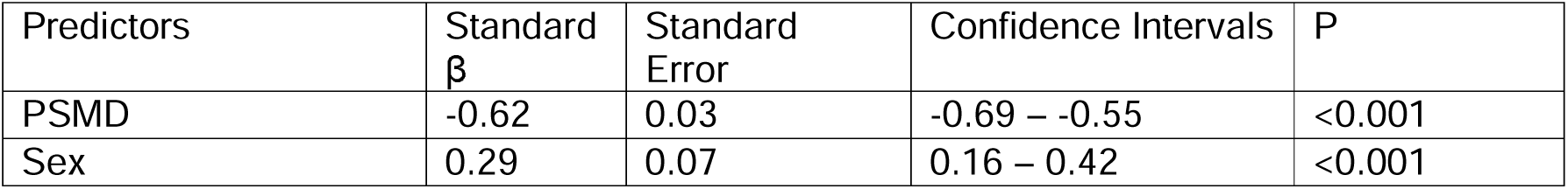
Results of Regression Model 2a (n = 579, DoF = 576, residual standard error = 0.73) with dependent variable Processing Speed. Significant effects (p<0.05) are shown in bold.

**Table S9.** Results of Regression Model 2b (n = 579, DoF = 575, residual standard error = 0.66) with dependent variable Processing Speed. Significant effects (p<0.05) are shown in bold.

| Predictors | Standard $\beta$ | Standard Error | Confidence Intervals | P |
| --- | --- | --- | --- | --- |
| PSMD | -0.24 | 0.05 | -0.33 – -0.15 | <0.001 |
| Sex | 0.18 | 0.06 | 0.06 – 0.30 | <0.01 |
| Age | -0.49 | 0.04 | -0.58 – -0.40 | <0.001 |

**Table S10.** Results of comparisons on Models 2a to 2c, using AIC and BIC, and the sum of squares derived from ANOVA comparisons.

|  | Difference in AIC | Difference in BIC | Difference in Sum Of Squares |
| --- | --- | --- | --- |
| Model 2a vs 2b | 97.08 | 92.72 | 58.50 |
| Model 2b vs 2c | -1.48 | -5.84 | 0.28 |

**Table S11.**
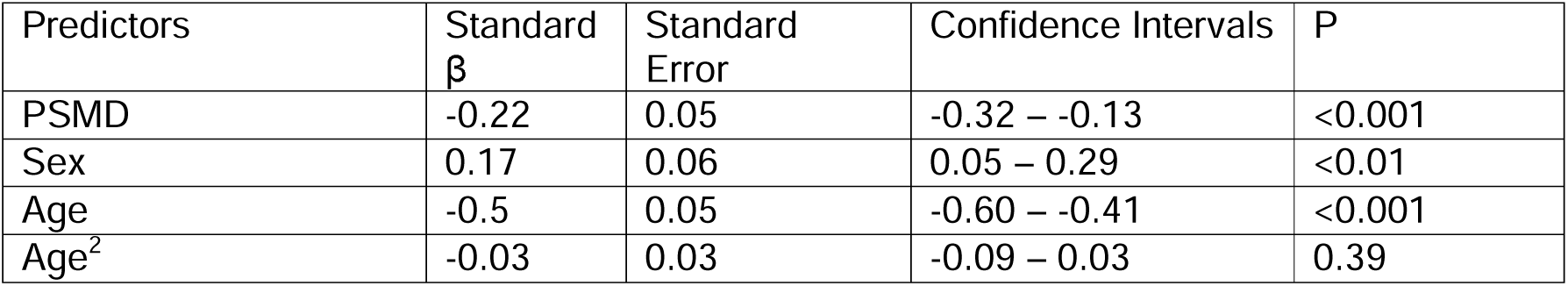
Results of Regression Model 2c (n = 579, DoF = 574, residual standard error = 0.66). Significant effects (p<0.05) are shown in bold.

| Predictors | Standard $\beta$ | Standard Error | Confidence Intervals | P |
| --- | --- | --- | --- | --- |
| PSMD | -0.22 | 0.05 | -0.32 – -0.13 | <0.001 |
| Sex | 0.17 | 0.06 | 0.05 – 0.29 | <0.01 |
| Age | -0.5 | 0.05 | -0.60 – -0.41 | <0.001 |
| Age<sup>2</sup> | -0.03 | 0.03 | -0.09 – 0.03 | 0.39 |

**Table S12.** Structural Equation Model 1a. Significant effects where confidence intervals do not cross zero are in bold.

| Dependent variable | Predictor variable | Label | Standard $\beta$ | Standard Error | z | p | Confidence Intervals |
| --- | --- | --- | --- | --- | --- | --- | --- |
| Speed | PP | c<sub>1</sub>' | -0.10 | 0.04 | -2.25 | 0.02 | -0.18 – -0.01 |
| Speed | PP<sup>2</sup> | c<sub>2</sub>' | -0.03 | 0.04 | -0.73 | 0.47 | -0.11 – 0.05 |
| PSMD | PP | a<sub>1</sub> | 0.42 | 0.04 | 11.15 | <0.01 | 0.35 – 0.49 |
| PSMD | PP<sup>2</sup> | a<sub>2</sub> | 0.11 | 0.04 | 3.07 | <0.01 | 0.04 – 0.18 |
| Speed | PSMD | b<sub>1</sub> | -0.54 | 0.04 | -12.53 | <0.01 | -0.63 - -0.46 |
| PSMD | Sex |  | 0.10 | 0.08 | 2.54 | 0.01 | 0.04 – 0.35 |
| PSMD | Head Motion |  | 0.16 | 0.04 | 3.68 | <0.01 | 0.10 – 0.27 |
| Speed | Sex |  | 0.13 | 0.07 | 3.66 | <0.01 | 0.12 – 0.41 |
| TAM of PP + PP<sup>2</sup> |  | (a<sub>1</sub> x b) + (a<sub>2</sub> x b) | 0.29 | 0.04 | 7.3 | <0.01 | 0.22 – 0.37 |

Abbreviations: PP, pulse pressure; PSMD, peak width of skeletonized mean diffusivity; TAM – Total Absolute Mediation.

**Table S13.** Structural Equation Model 1b. Significant effects where confidence intervals do not cross zero are in bold.

| Dependent variable | Predictor variable | Label | Standard $\beta$ | Standard Error | z | p | Confidence Intervals |
| --- | --- | --- | --- | --- | --- | --- | --- |
| Speed | PP | c<sub>1</sub>' | 0.01 | 0.04 | 0.15 | 0.89 | -0.07 – 0.09 |
| Speed | PP<sup>2</sup> | c<sub>2</sub>' | -0.04 | 0.04 | -0.92 | 0.36 | -0.11 – 0.04 |
| PSMD | PP | a<sub>1</sub> | 0.42 | 0.04 | 11.15 | <0.01 | 0.35 – 0.49 |
| PSMD | PP<sup>2</sup> | a<sub>2</sub> | 0.11 | 0.04 | 3.07 | <0.01 | 0.04 – 0.18 |
| Speed | PSMD | b<sub>1</sub> | -0.20 | 0.06 | -3.40 | <0.01 | -0.30 – -0.08 |
| PSMD | Sex |  | 0.10 | 0.08 | 2.54 | 0.01 | 0.04 – 0.35 |
| PSMD | Head Motion |  | 0.16 | 0.04 | 3.68 | <0.01 | 0.10 – 0.27 |
| Speed | Sex |  | 0.13 | 0.07 | 1.93 | 0.05 | -0.00 – 0.26 |
| Speed | Age |  | -0.54 | 0.05 | -10.00 | <0.01 | -0.61 – -0.41 |
| TAM of PP + PP<sup>2</sup> |  | (a<sub>1</sub> x b) + (a<sub>2</sub> x b) | 0.11 | 0.03 | 3.05 | <0.01 | 0.04 – 0.17 |

Abbreviations: PP, pulse pressure; PSMD, peak width of skeletonized mean diffusivity; TAM – Total Absolute Mediation.

**Table S14.** Likelihood ratio test results on Structural Equation Model 1b, comparing the full model to a version where the path to Age was essentially removed, by constraining it to be equal to zero.

| Model | DF | AIC | BIC | Chi-sq | p |
| --- | --- | --- | --- | --- | --- |
| SEM 1b, full model | 2 | 9, 889 | 10, 002 | 371.96 |  |
| SEM 1b, constraining Age | 3 | 9, 982 | 10, 091 | 467.14 | <0.001 |

Abbreviations: AIC, Akaike Information Criterion; BIC, Bayesian Information Criterion; DF, degrees of freedom.

**Table S15.** Structural Equation Model 1c. Significant effects where confidence intervals do not cross zero are in bold.

| Dependent variable | Predictor variable | label | Standard $\beta$ | Standard Error | z | p | Confidence intervals |
| --- | --- | --- | --- | --- | --- | --- | --- |
| Speed | PP | c<sub>1</sub>' | 0.01 | 0.04 | 0.30 | 0.77 | -0.07 – 0.10 |
| Speed | PP<sup>2</sup> | c<sub>2</sub>' | -0.03 | 0.04 | -0.89 | 0.38 | -0.11 – 0.04 |
| PSMD | PP | $a_1$ | 0.42 | 0.04 | 11.15 | <0.01 | 0.35 – 0.49 |
| PSMD | PP<sup>2</sup> | $a_2$ | 0.11 | 0.04 | 3.07 | <0.01 | 0.04 – 0.18 |
| Speed | PSMD | $b_1$ | -0.19 | 0.06 | -3.00 | <0.01 | -0.29 – -0.06 |
| PSMD | Sex |  | 0.10 | 0.08 | 2.54 | 0.01 | 0.04 – 0.35 |
| PSMD | Head Motion |  | 0.16 | 0.04 | 3.68 | <0.01 | 0.10 – 0.27 |
| Speed | Sex |  | 0.07 | 0.07 | 1.74 | 0.08 | -0.01 – 0.26 |
| Speed | Age |  | -0.55 | 0.06 | -9.50 | <0.01 | -0.63 – -0.42 |
| Speed | Age<sup>2</sup> |  | -0.03 | 0.03 | -0.80 | 0.42 | -0.09 – 0.04 |
| TAM of PP + PP<sup>2</sup> | | $(a_1 \times b) + (a_2 \times b)$ | 0.10 | 0.03 | 2.73 | 0.01 | 0.03 – 0.16 |

Abbreviations: PP, pulse pressure; PSMD, peak width of skeletonized mean diffusivity; TAM – Total Absolute Mediation.

**Table S16.**
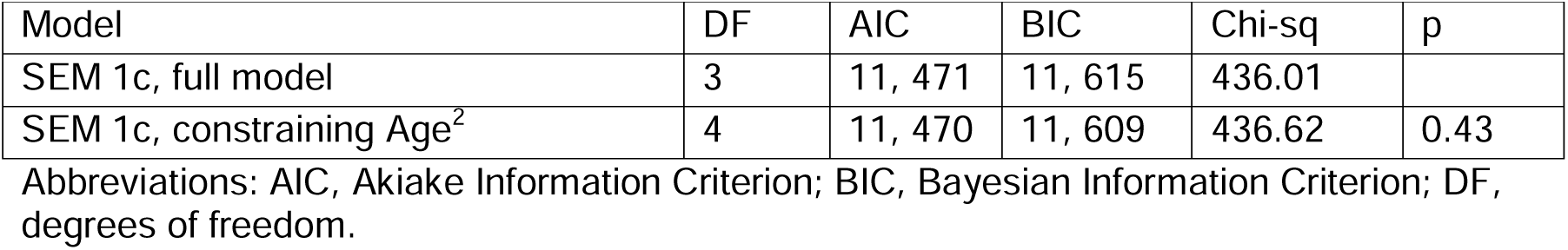
Likelihood ratio test results comparing Structural Equation Model 1c, comparing the full model to a version where the path to Age^2^ was essentially removed, by constraining it to be equal to zero.

| Model | DF | AIC | BIC | Chi-sq | p |
| --- | --- | --- | --- | --- | --- |
| SEM 1c, full model | 3 | 11, 471 | 11, 615 | 436.01 |  |
| SEM 1c, constraining Age<sup>2</sup> | 4 | 11, 470 | 11, 609 | 436.62 | 0.43 |

Abbreviations: AIC, Akaike Information Criterion; BIC, Bayesian Information Criterion; DF, degrees of freedom.

**Table S17.** Structural Equation Model 1d. Significant effects where confidence intervals do not cross zero are in bold.

| Dependent variable | Predictor variable | label | Standard $\beta$ | Standard Error | z | p | Confidence Intervals |
| --- | --- | --- | --- | --- | --- | --- | --- |
| Speed | SSBP | $c_{1'}$ | 0.03 | 0.04 | 0.73 | 0.47 | -0.05 – 0.10 |
| Speed | SSBPP<sup>2</sup> | $c_{2'}$ | 0.01 | 0.03 | 0.40 | 0.69 | -0.05 – 0.08 |
| Speed | PP | $c_{3'}$ | 0.00 | 0.04 | -0.07 | 0.95 | -0.09 – 0.08 |
| Speed | PP<sup>2</sup> | $c_{4'}$ | -0.04 | 0.04 | -1.01 | 0.31 | -0.12 – 0.03 |
| Speed | HRV | $c_{5'}$ | 0.07 | 0.05 | 1.47 | 0.14 | -0.02 – 0.16 |
| Speed | HRV<sup>2</sup> | $c_{6'}$ | -0.01 | 0.04 | -0.21 | 0.84 | -0.07 – 0.08 |
| PSMD | SSBP | $a_1$ | -0.02 | 0.05 | -0.43 | 0.66 | -0.11 – 0.07 |
| PSMD | SSBPP<sup>2</sup> | $a_2$ | 0.00 | 0.04 | -0.04 | 0.97 | -0.08 – 0.07 |
| PSMD | PP | $a_3$ | 0.31 | 0.04 | 6.90 | <0.01 | 0.22 – 0.40 |
| PSMD | PP<sup>2</sup> | $a_4$ | 0.09 | 0.04 | 2.42 | 0.02 | 0.02 – 0.16 |
| PSMD | HRV | $a_5$ | -0.35 | 0.04 | -8.78 | <0.01 | -0.44 – -0.28 |
| PSMD | HRV<sup>2</sup> | a<sub>6</sub> | -0.01 | 0.04 | -0.33 | 0.74 | -0.08 – 0.07 |
| Speed | PSMD | b<sub>1</sub> | -0.20 | 0.06 | -3.37 | <0.01 | -0.30 – -0.08 |
| PSMD | Sex |  | 0.12 | 0.07 | 3.27 | <0.01 | 0.09 – 0.39 |
| PSMD | Head Motion |  | 0.13 | 0.04 | 3.16 | <0.01 | 0.07 – 0.24 |
| Speed | Sex |  | 0.07 | 0.07 | 1.75 | 0.08 | -0.01 – 0.27 |
| Speed | Age |  | -0.49 | 0.06 | -8.16 | <0.01 | -0.59 – -0.36 |
| Total Absolute Mediation of SSBP + SSBP<sup>2</sup> |  | (a<sub>1</sub> x b) + (a<sub>2</sub> x b) | 0.00 | 0.01 | 0.48 | 0.63 | 0.00 – 0.04 |
| TAM of PP + PP<sup>2</sup> |  | (a<sub>3</sub> x b) + (a<sub>4</sub> x b) | 0.08 | 0.03 | 2.91 | <0.01 | 0.03 – 0.13 |
| TAM of HRV + HRV<sup>2</sup> |  | (a<sub>5</sub> x b) + (a<sub>6</sub> x b) | 0.07 | 0.02 | 2.90 | <0.01 | 0.03 – 0.12 |

Abbreviations: HRV, heart rate variability; SSBP, steady state blood pressure; PP, pulse pressure; PSMD, peak width of skeletonized mean diffusivity; TAM – Total Absolute Mediation.

**Table S18.** Structural Equation Model 1a using observed variables of pulse pressure. Significant effects where confidence intervals do not cross zero are in bold.

| Dependent variable | Predictor variable | Label | Standard $\beta$ | Standard Error | z | p | Confidence Intervals |
| --- | --- | --- | --- | --- | --- | --- | --- |
| Speed | PP | c<sub>1</sub>' | -0.107 | 0.045 | -2.36 | 0.02 | -0.19 – -0.02 |
| Speed | PP<sup>2</sup> | c<sub>2</sub>' | -0.018 | 0.043 | -0.431 | 0.67 | -0.10 – 0.07 |
| PSMD | PP | a<sub>1</sub> | 0.449 | 0.04 | 11.23 | <0.01 | 0.37 – 0.53 |
| PSMD | PP<sup>2</sup> | a<sub>2</sub> | 0.111 | 0.038 | 2.922 | <0.01 | 0.04 – 0.19 |
| Speed | PSMD | b<sub>1</sub> | -0.549 | 0.049 | -11.291 | <0.01 | -0.63 – -0.46 |
| PSMD | Sex |  | 0.139 | 0.08 | 1.744 | 0.08 | -0.02 – 0.30 |
| PSMD | Head Motion |  | 0.145 | 0.047 | 3.077 | <0.01 | 0.08 – 0.26 |
| Speed | Sex |  | -0.107 | 0.045 | -2.36 | 0.02 | 0.19 – 0.48 |
| TAM of PP + PP<sup>2</sup> |  | (a<sub>1</sub> x b) + (a<sub>2</sub> x b) | 0.31 | 0.04 | 7.3 | <0.01 | 0.23 – 0.40 |

Abbreviations: PP, pulse pressure; PSMD, peak width of skeletonized mean diffusivity; TAM – Total Absolute Mediation.

**Table S19.** Structural Equation Model 1b using observed variables of pulse pressure. Significant effects where confidence intervals do not cross zero are in bold.

| Dependent | Predictor | Label | Standard | Standard | z | p | Confidence |
| --- | --- | --- | --- | --- | --- | --- | --- |

| variable | variable | | $\beta$ | Error | | | Intervals |
| --- | --- | --- | --- | --- | --- | --- | --- |
| Speed | PP | $c_{1'}$ | 0.006 | 0.042 | 0.148 | 0.88 | -0.08 – 0.09 |
| Speed | PP<sup>2</sup> | $c_{2'}$ | -0.008 | 0.038 | -0.204 | 0.84 | -0.08 – 0.07 |
| PSMD | PP | $a_1$ | 0.449 | 0.04 | 11.229 | <0.01 | 0.37 – 0.53 |
| PSMD | PP<sup>2</sup> | $a_2$ | 0.111 | 0.038 | 2.923 | <0.01 | 0.04 – 0.19 |
| Speed | PSMD | $b_1$ | -0.172 | 0.064 | -2.706 | <0.01 | -0.29 – -0.05 |
| PSMD | Sex |  | 0.139 | 0.08 | 1.743 | 0.08 | -0.02 – 0.30 |
| PSMD | Head Motion |  | 0.145 | 0.047 | 3.078 | <0.01 | 0.08 – 0.27 |
| Speed | Sex |  | 0.196 | 0.071 | 2.769 | <0.01 | 0.06 – 0.34 |
| Speed | Age |  | -0.552 | 0.056 | -9.848 | <0.01 | -0.67 – -0.45 |
| TAM of PP + PP<sup>2</sup> | | $(a_1 \times b) + (a_2 \times b)$ | 0.10 | 0.04 | 2.51 | <0.01 | 0.03 – 0.18 |

Abbreviations: PP, pulse pressure; PSMD, peak width of skeletonized mean diffusivity; TAM – Total Absolute Mediation.

**Table S20.** Likelihood ratio test results on Structural Equation Model 1b using observed variables, comparing the full model to a version where the path to Age was essentially removed, by constraining it to be equal to zero.

| Model | DF | AIC | BIC | Chi-sq | p |
| --- | --- | --- | --- | --- | --- |
| SEM 1b, full model | 2 | 8, 678 | 8, 789 | 377.65 |  |
| SEM 1b, constraining Age | 3 | 8, 777 | 8, 882 | 427.22 | <0.001 |

Abbreviations: AIC, Akaike Information Criterion; BIC, Bayesian Information Criterion; DF, degrees of freedom.

**Table S21.**
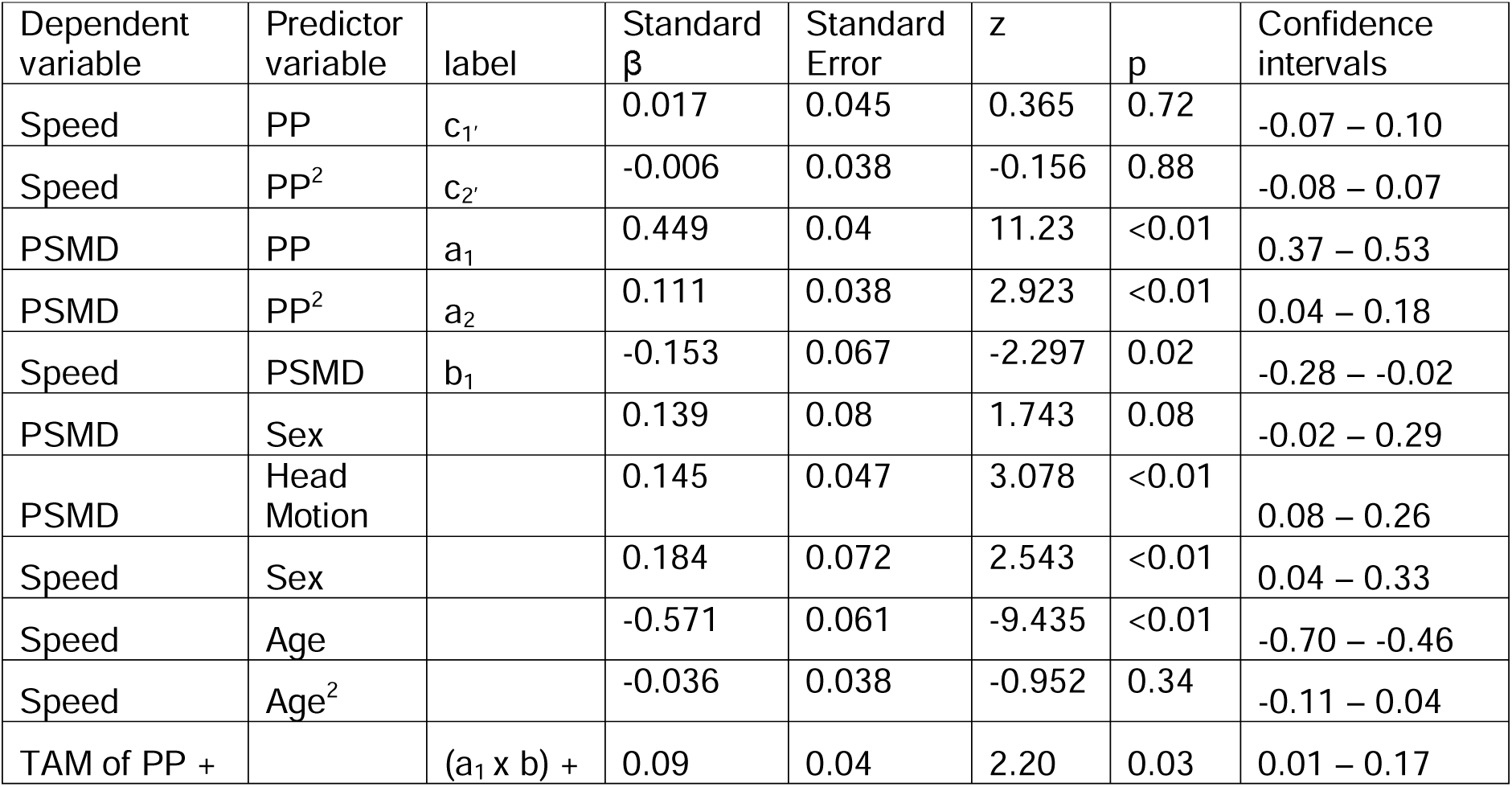
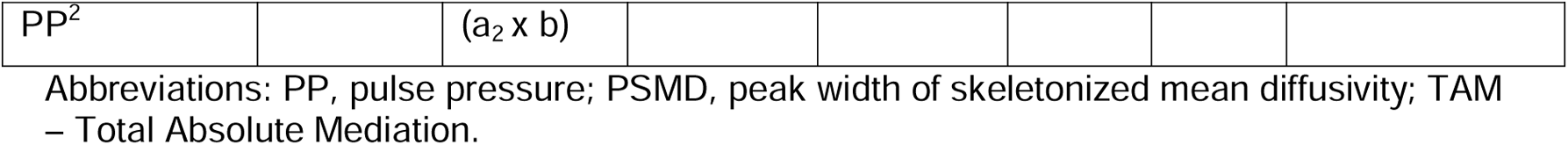
Structural Equation Model 1c using observed variables of pulse pressure. Significant effects where confidence intervals do not cross zero are in bold.

**Table S22.**
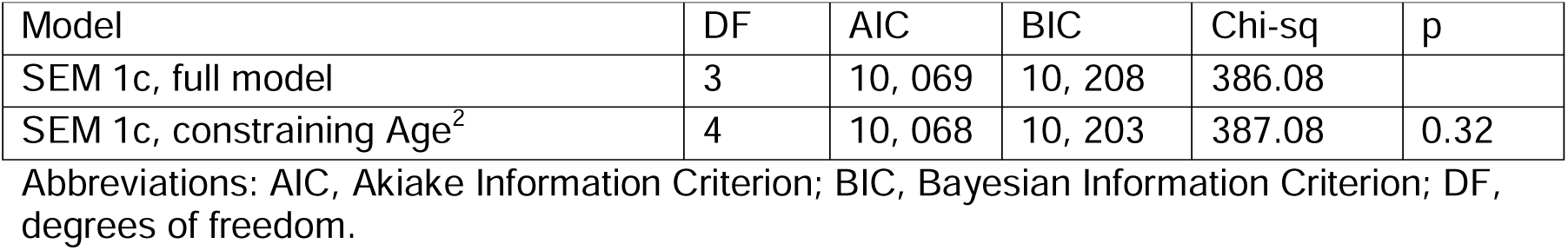
Likelihood ratio test results comparing Structural Equation Model 1c using observed variables of pulse pressure, comparing the full model to a version where the path to Age^2^ was essentially removed, by constraining it to be equal to zero.

| Model | DF | AIC | BIC | Chi-sq | p |
| --- | --- | --- | --- | --- | --- |
| SEM 1c, full model | 3 | 10, 069 | 10, 208 | 386.08 |  |
| SEM 1c, constraining Age<sup>2</sup> | 4 | 10, 068 | 10, 203 | 387.08 | 0.32 |

Abbreviations: AIC, Akaike Information Criterion; BIC, Bayesian Information Criterion; DF, degrees of freedom.

**Table S23.**
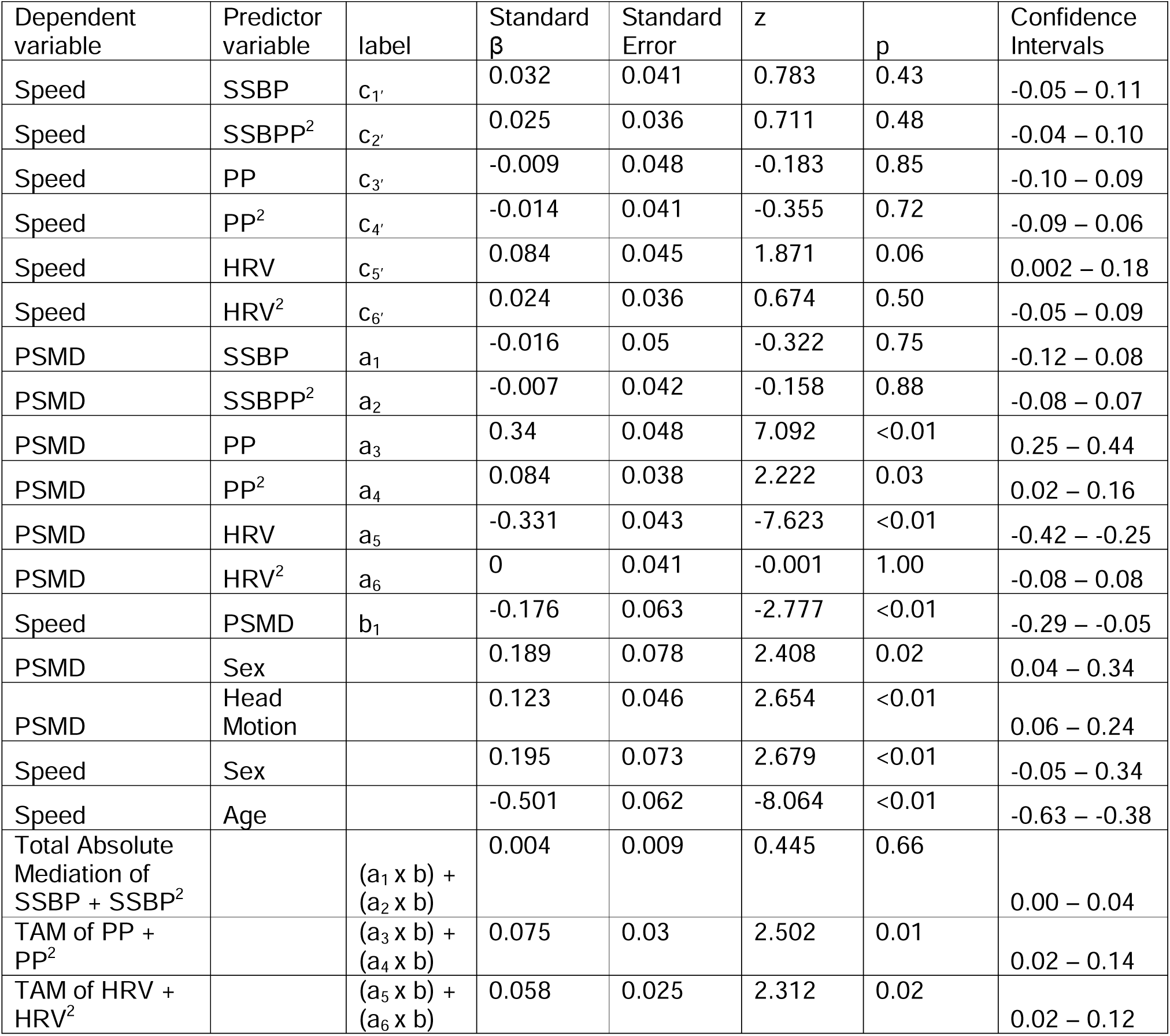
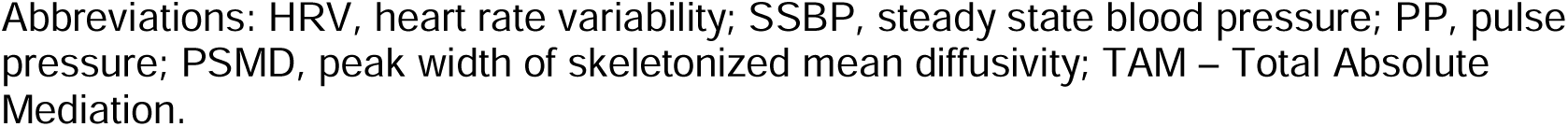
Structural Equation Model 1d using observed variables of pulse pressure and steady state blood pressure. Significant effects where confidence intervals do not cross zero are in bold

| Dependent variable | Predictor variable | label | Standard $\beta$ | Standard Error | z | p | Confidence Intervals |
| --- | --- | --- | --- | --- | --- | --- | --- |
| Speed | SSBP | c<sub>1'</sub> | 0.032 | 0.041 | 0.783 | 0.43 | -0.05 – 0.11 |
| Speed | SSBPP<sup>2</sup> | c<sub>2'</sub> | 0.025 | 0.036 | 0.711 | 0.48 | -0.04 – 0.10 |
| Speed | PP | c<sub>3'</sub> | -0.009 | 0.048 | -0.183 | 0.85 | -0.10 – 0.09 |
| Speed | PP<sup>2</sup> | c<sub>4'</sub> | -0.014 | 0.041 | -0.355 | 0.72 | -0.09 – 0.06 |
| Speed | HRV | c<sub>5'</sub> | 0.084 | 0.045 | 1.871 | 0.06 | 0.002 – 0.18 |
| Speed | HRV<sup>2</sup> | c<sub>6'</sub> | 0.024 | 0.036 | 0.674 | 0.50 | -0.05 – 0.09 |
| PSMD | SSBP | a<sub>1</sub> | -0.016 | 0.05 | -0.322 | 0.75 | -0.12 – 0.08 |
| PSMD | SSBPP<sup>2</sup> | a<sub>2</sub> | -0.007 | 0.042 | -0.158 | 0.88 | -0.08 – 0.07 |
| PSMD | PP | a<sub>3</sub> | 0.34 | 0.048 | 7.092 | <0.01 | 0.25 – 0.44 |
| PSMD | PP<sup>2</sup> | a<sub>4</sub> | 0.084 | 0.038 | 2.222 | 0.03 | 0.02 – 0.16 |
| PSMD | HRV | a<sub>5</sub> | -0.331 | 0.043 | -7.623 | <0.01 | -0.42 – -0.25 |
| PSMD | HRV<sup>2</sup> | a<sub>6</sub> | 0 | 0.041 | -0.001 | 1.00 | -0.08 – 0.08 |
| Speed | PSMD | b<sub>1</sub> | -0.176 | 0.063 | -2.777 | <0.01 | -0.29 – -0.05 |
| PSMD | Sex |  | 0.189 | 0.078 | 2.408 | 0.02 | 0.04 – 0.34 |
| PSMD | Head Motion |  | 0.123 | 0.046 | 2.654 | <0.01 | 0.06 – 0.24 |
| Speed | Sex |  | 0.195 | 0.073 | 2.679 | <0.01 | -0.05 – 0.34 |
| Speed | Age |  | -0.501 | 0.062 | -8.064 | <0.01 | -0.63 – -0.38 |
| Total Absolute Mediation of SSBP + SSBP<sup>2</sup> |  | (a<sub>1</sub> x b) + (a<sub>2</sub> x b) | 0.004 | 0.009 | 0.445 | 0.66 | 0.00 – 0.04 |
| TAM of PP + PP<sup>2</sup> |  | (a<sub>3</sub> x b) + (a<sub>4</sub> x b) | 0.075 | 0.03 | 2.502 | 0.01 | 0.02 – 0.14 |
| TAM of HRV + HRV<sup>2</sup> |  | (a<sub>5</sub> x b) + (a<sub>6</sub> x b) | 0.058 | 0.025 | 2.312 | 0.02 | 0.02 – 0.12 |

Abbreviations: HRV, heart rate variability; SSBP, steady state blood pressure; PP, pulse pressure; PSMD, peak width of skeletonized mean diffusivity; TAM – Total Absolute Mediation.

**Table S24.** Likelihood ratio test results comparing Structural Equation Model 2a, where no paths vary with Age, to Structural Equation Model 2b, where paths ‘a1’ and ‘a2’ vary across three age groups: Young (18-44 years, n=190), Middle (44-65 years, n=190) and Old (65-87 years, n=190).

| Model | DF | AIC | BIC | Chi-sq | p |
| --- | --- | --- | --- | --- | --- |
| SEM 2a: no paths vary across Age groups | 13 | 7, 062 | 7, 279 | 10.19 |  |
| SEM 2b: 'a<sub>1</sub>' and 'a<sub>2</sub>' vary across Age groups | 9 | 7, 068 | 7, 302 | 7.82 | 0.67 |

Abbreviations: AIC, Akiake Information Criterion; BIC, Bayesian Information Criterion; DF, degrees of freedom.

**Table S25.** Likelihood ratio test results on Structural Equation Model 3a, comparing the full model to a version where the ‘b_2_’ path between processing speed and the ability discrepancy was essentially removed, by constraining it to be equal to zero.

| Model | DF | AIC | BIC | Chi-sq | p |
| --- | --- | --- | --- | --- | --- |
| SEM 3a, full model | 6 | 11, 019 | 11, 149 | 366 |  |
| SEM 3a, constraining 'b<sub>2</sub>' | 7 | 11, 021 | 11, 147 | 370 | 0.04 |

Abbreviations: AIC, Akiake Information Criterion; BIC, Bayesian Information Criterion; DF, degrees of freedom.

**Table S26.**
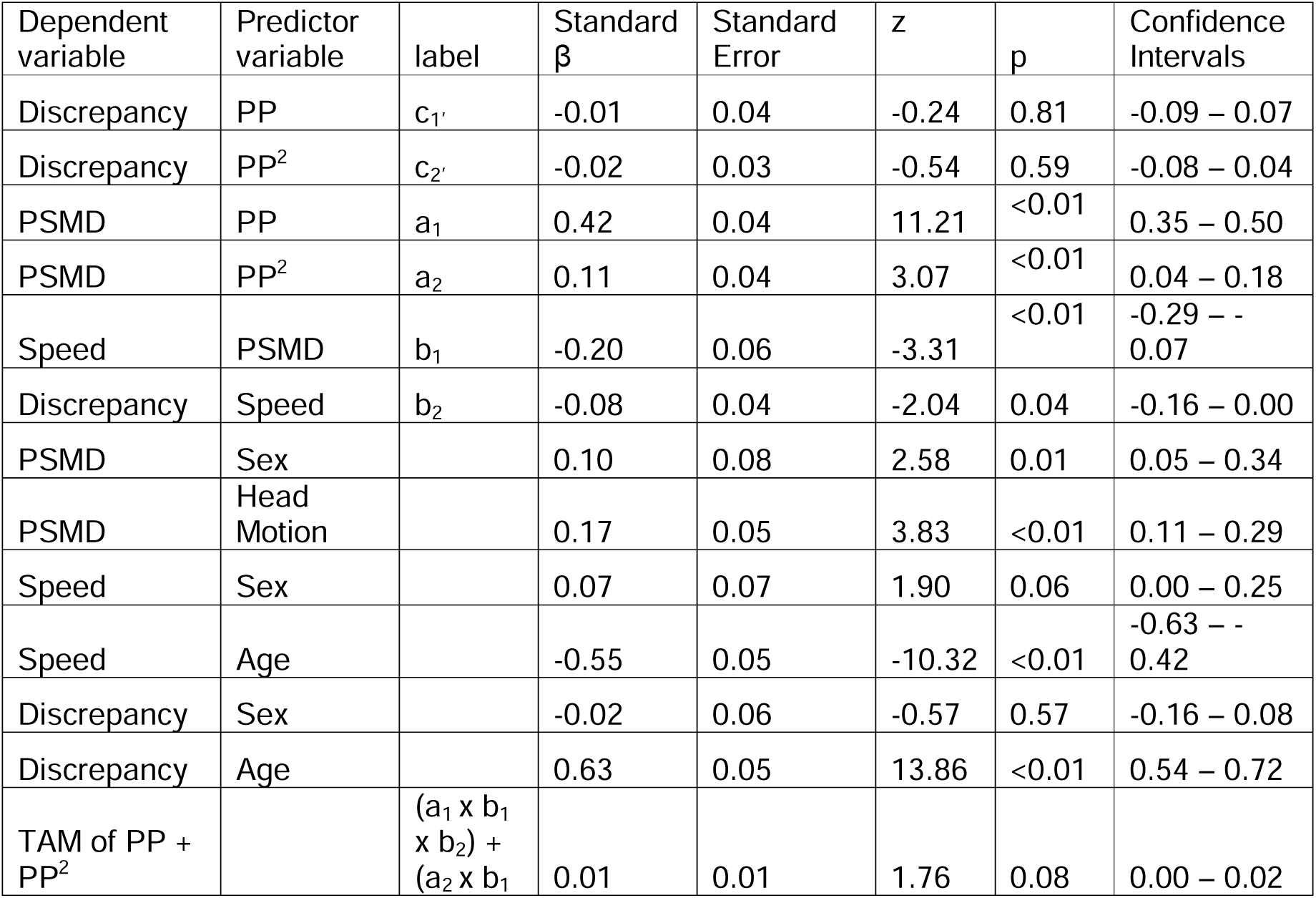
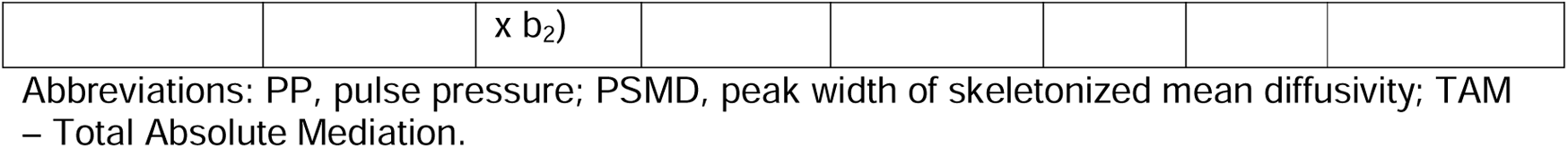
Structural Equation Model 3a. Significant effects where confidence intervals do not cross zero are in bold.

**Table S27.**
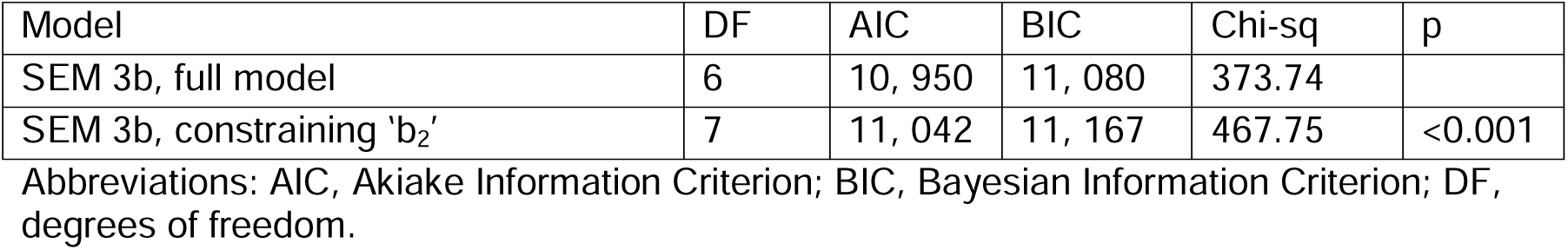
Likelihood ratio test results on Structural Equation Model 3b, comparing the full model to a version where the ‘b_2_’ path between processing speed and fluid intelligence was essentially removed, by constraining it to be equal to zero.

**Table S28.** Structural Equation Model 3b. Significant effects where confidence intervals do not cross zero are in bold.

| Dependent variable | Predictor variable | label | Standard $\beta$ | Standard Error | z | p | Confidence Intervals |
| --- | --- | --- | --- | --- | --- | --- | --- |
| Fluid | PP | c<sub>1</sub>' | -0.06 | 0.03 | -1.71 | 0.09 | -0.13 – 0.01 |
| Fluid | PP<sup>2</sup> | c<sub>2</sub>' | 0.03 | 0.03 | 0.94 | 0.35 | -0.03 – 0.08 |
| PSMD | PP | a<sub>1</sub> | 0.42 | 0.04 | 11.21 | <0.01 | 0.35 – 0.50 |
| PSMD | PP<sup>2</sup> | a<sub>2</sub> | 0.11 | 0.04 | 3.07 | <0.01 | 0.04 – 0.18 |
| Speed | PSMD | b<sub>1</sub> | -0.20 | 0.06 | -3.31 | <0.01 | -0.29 – -0.07 |
| Fluid | Speed | b<sub>2</sub> | 0.38 | 0.04 | 9.12 | <0.01 | 0.31 – 0.48 |
| PSMD | Sex |  | 0.10 | 0.08 | 2.58 | 0.01 | 0.05 – 0.34 |
| PSMD | Head Motion |  | 0.17 | 0.05 | 3.83 | <0.01 | 0.11 – 0.29 |
| Speed | Sex |  | 0.07 | 0.07 | 1.90 | 0.06 | 0.00 – 0.25 |
| Speed | Age |  | -0.55 | 0.05 | -10.32 | <0.01 | -0.63 – -0.42 |
| Fluid | Sex |  | 0.07 | 0.06 | 2.41 | 0.02 | 0.02 – 0.26 |
| Fluid | Age |  | -0.38 | 0.04 | -8.75 | <0.01 | -0.46 – -0.29 |
| TAM of PP + PP<sup>2</sup> |  | (a<sub>1</sub> x b<sub>1</sub> x b<sub>2</sub>) + (a<sub>2</sub> x b<sub>1</sub> x b<sub>2</sub>) | 0.04 | 0.01 | 2.77 | 0.01 | 0.01 – 0.07 |

Abbreviations: PP, pulse pressure; PSMD, peak width of skeletonized mean diffusivity; TAM – Total Absolute Mediation.

**Table S29.** Likelihood ratio test results comparing Structural Equation Model 3B with the expected and reversed orders of variables, excluding covariats of no interest. No p-value is given when models have the same degrees of freedom.

| Model | DF | AIC | BIC | Chi-sq |
| --- | --- | --- | --- | --- |
| Expected order:<br>Speed (b<sub>2</sub>) – Fluid | 3 | 7, 328 | 7, 380 | 43.59 |
| Reversed order:<br>Fluid (b<sub>2</sub>) – Speed | 3 | 7, 346 | 7, 398 | 62.39 |

Abbreviations:
AIC, Akiake Information Criterion; BIC, Bayesian Information Criterion; DF, degrees of freedom; PP, pulse pressure; PSMD, peak width of skeletonized mean diffusivity.

**Table S30.**
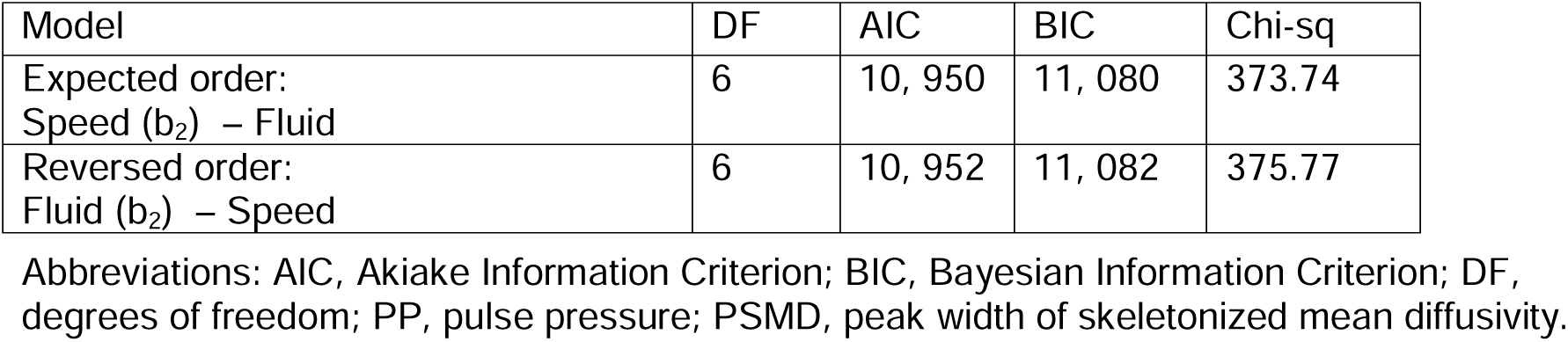
Likelihood ratio test results comparing Structural Equation Model 3B with the expected and reversed orders of variables, including covariates of no interest. No p-value is given when models have the same degrees of freedom.

| Model | DF | AIC | BIC | Chi-sq |
| --- | --- | --- | --- | --- |
| Expected order:<br>Speed (b<sub>2</sub>) – Fluid | 6 | 10, 950 | 11, 080 | 373.74 |
| Reversed order:<br>Fluid (b<sub>2</sub>) – Speed | 6 | 10, 952 | 11, 082 | 375.77 |

Abbreviations: AIC, Akiake Information Criterion; BIC, Bayesian Information Criterion; DF, degrees of freedom; PP, pulse pressure; PSMD, peak width of skeletonized mean diffusivity.

### Author contributions

Designed research: D.L.O.K., R.N.A.H., K.A.T., J.B.R.

Performed research: D.L.O.K., R.N.A.H., K.A.T.

Analysed data: D.L.O.K., R.N.A.H., M.C., K.A.T.

Wrote the paper: D.L.O.K., R.N.A.H., M.C., J.B.R, K.A.T.

### Funders

Cam-CAN was supported by the Biotechnology and Biological Sciences Research Council Grant BB/H008217/1.

D.L.O.K. was supported by a Doctoral Training Programme studentship awarded by the Biotechnology and Biological Sciences Research Council (BBSRC BB/M011194/1)

K.A.T. was supported by Fellowship awards from the Guarantors of Brain (G101149) and Alzheimer’s Society, UK (grant number 602).

R.N.A.H. was supported by the UK Medical Research Council (SUAG/046 G101400) and the European Union’s Horizon 2020 research and innovation programme (‘LifeBrain’, Grant Agreement No. 732592).

J.B.R. is funded by the Welcome Trust (220258), the Cambridge University Centre for Frontotemporal Dementia, the Medical Research Council (MC_UU_00030/14; MR/T033371/1); the National Institute for Health Research Cambridge Biomedical Research Centre (NIHR203312: BRC-1215-20014) and the Holt Fellowship. The views expressed are those of the authors and not necessarily those of the NIHR or the Department of Health and Social Care.

We thank the Cam-CAN respondents and their primary care teams in Cambridge for their participation in this study, and colleagues at the MRC Cognition and Brain Sciences Unit MEG and MRI facilities for their assistance. Further information about the Cam-CAN corporate authorship membership can be found at https://cam-can.mrc-cbu.cam.ac.uk/corpauth/#14 (**list #14**).

For the purpose of open access, the author has applied a Creative Commons Attribution (CC BY) licence to any Author Accepted Manuscript version arising from this submission.

## Data Availability

The raw data are available on request from https://camcan-archive.mrc-cbu.cam.ac.uk/dataaccess/. Statistical analyses were performed in R (version 4.0.2) and R-Studio 50. PSMD was calculated using the release 1.8.2 (https://github.com/miac-research/psmd/releases) 12. CSVs for the summary measures and R code for regression models and SEMS are available at: https://github.com/DebsKing/Pulse_pressure_impairs_cognition_via_white_matter_disruption.git.

https://camcan-archive.mrc-cbu.cam.ac.uk/dataaccess/

## Notes

### Competing Interest Statement

The authors have declared no competing interest.

### Author Declarations

The study was approved by the Cambridgeshire 2 (now East of England, Cambridge Central) Research Ethics Committee (reference: 10/H0308/50).

## BIBLIOGRAPHY

1. Thorin-Trescases N, de Montgolfier O, Pinçon A, Raignault A, Caland L, Labbé P, Thorin E. Impact of pulse pressure on cerebrovascular events leading to age-related cognitive decline. American Journal of Physiology-Heart and Circulatory Physiology. 2018;314:H1214–H1224.

2. Black S, Gao F, Bilbao J. Understanding White Matter Disease: Imaging-Pathological Correlations in Vascular Cognitive Impairment. Stroke [Internet]. 2009 [cited 2023 Apr 19];40. Available from: https://www.ahajournals.org/doi/10.1161/STROKEAHA.108.537704

3. Rowbotham GF, Little E. Circulations of the cerebral hemispheres. British Journal of Surgery. 2005;52:8–21.

4. Kang P, Ying C, Chen Y, Ford AL, An H, Lee J-M. Oxygen Metabolic Stress and White Matter Injury in Patients With Cerebral Small Vessel Disease. Stroke. 2022;53:1570– 1579.

5. Pantoni L, Garcia JH, Gutierrez JA. Cerebral White Matter Is Highly Vulnerable to Ischemia. Stroke. 1996;27:1641–1647.

6. Martinez Sosa S, Smith KJ. Understanding a role for hypoxia in lesion formation and location in the deep and periventricular white matter in small vessel disease and multiple sclerosis. Clinical Science. 2017;131:2503–2524.

7. Kennedy KM, Raz N. Pattern of normal age-related regional differences in white matter microstructure is modified by vascular risk. Brain Research. 2009;1297:41–56.

8. Reas Emilie T., Laughlin Gail A., Hagler Donald J., Lee Roland R., Dale Anders M., McEvoy Linda K. Age and Sex Differences in the Associations of Pulse Pressure With White Matter and Subcortical Microstructure. Hypertension. 0:HYPERTENSIONAHA.120.16446.

9. Wartolowska KA, Webb AJ. White matter damage due to pulsatile versus steady blood pressure differs by vascular territory: A cross-sectional analysis of the UK Biobank cohort study. J Cereb Blood Flow Metab. 2022;42:802–810.

10. Wei J, Palta P, Meyer ML, Kucharska-Newton A, Pence BW, Aiello AE, Power MC, Walker KA, Sharrett AR, Tanaka H, et al. Aortic Stiffness and White Matter Microstructural Integrity Assessed by Diffusion Tensor Imaging: The ARIC- NCS. JAHA. 2020;9:e014868.

11. King DLO, Henson RN, Kievit R, Wolpe N, Brayne C, Tyler LK, Rowe JB, Cam-CAN, Bullmore ET, Calder AC, et al. Distinct components of cardiovascular health are linked with age-related differences in cognitive abilities. Sci Rep. 2023;13:978.

12. Baykara E, Gesierich B, Adam R, Tuladhar AM, Biesbroek JM, Koek HL, Ropele S, Jouvent E, Chabriat H, Ertl-Wagner B, et al. A Novel Imaging Marker for Small Vessel Disease Based on Skeletonization of White Matter Tracts and Diffusion Histograms. Annals of Neurology. 2016;80:581–592.

13. Beaudet G, Tsuchida A, Petit L, Tzourio C, Caspers S, Schreiber J, Pausova Z, Patel Y, Paus T, Schmidt R, et al. Age-Related Changes of Peak Width Skeletonized Mean Diffusivity (PSMD) Across the Adult Lifespan: A Multi-Cohort Study. Front. Psychiatry. 2020;11:342.

14. Beaudet G, Tsuchida A, Petit L, Tzourio C, Caspers S, Schreiber J, Pausova Z, Patel Y, Paus T, Schmidt R, et al. Age-Related Changes of Peak Width Skeletonized Mean Diffusivity (PSMD) Across the Adult Lifespan: A Multi-Cohort Study. Front Psychiatry [Internet]. 2020 [cited 2021 Mar 2];11. Available from: https://www.ncbi.nlm.nih.gov/pmc/articles/PMC7212692/

15. Zanon Zotin MC, Yilmaz P, Sveikata L, Schoemaker D, van Veluw SJ, Etherton MR, Charidimou A, Greenberg SM, Duering M, Viswanathan A. Peak Width of Skeletonized Mean Diffusivity: A Neuroimaging Marker for White Matter Injury. Radiology. 2023;306:e212780.

16. Low A, Mak E, Stefaniak JD, Malpetti M, Nicastro N, Savulich G, Chouliaras L, Markus HS, Rowe JB, O’Brien JT. Peak Width of Skeletonized Mean Diffusivity as a Marker of Diffuse Cerebrovascular Damage. Front. Neurosci. 2020;14:238.

17. Lam BYK, Leung KT, Yiu B, Zhao L, Biesbroek JM, Au L, Tang Y, Wang K, Fan Y, Fu J-H, et al. Peak width of skeletonized mean diffusivity and its association with age-related cognitive alterations and vascular risk factors. Alzheimers Dement (Amst). 2019;11:721–729.

18. Koncz R, Wen W, Makkar SR, Lam BCP, Crawford JD, Rowe CC, Sachdev P. The Interaction Between Vascular Risk Factors, Cerebral Small Vessel Disease, and Amyloid Burden in Older Adults. JAD. 2022;86:1617–1628.

19. King DLO, Henson RN, Kievit R, Wolpe N, Brayne C, Tyler LK, Rowe JB, Cam-CAN, Bullmore ET, Calder AC, et al. Distinct components of cardiovascular health are linked with age-related differences in cognitive abilities. Sci Rep. 2023;13:978.

20. Filley CM, Fields RD. White matter and cognition: making the connection. Journal of Neurophysiology. 2016;116:2093–2104.

21. Deary IJ, Ritchie SJ, Muñoz Maniega S, Cox SR, Valdés Hernández MC, Luciano M, Starr JM, Wardlaw JM, Bastin ME. Brain Peak Width of Skeletonized Mean Diffusivity (PSMD) and Cognitive Function in Later Life. Front Psychiatry. 2019;10:524.

22. Oberlin LE, Verstynen TD, Burzynska AZ, Voss MW, Prakash RS, Chaddock-Heyman L, Wong C, Fanning J, Awick E, Gothe N, et al. White matter microstructure mediates the relationship between cardiorespiratory fitness and spatial working memory in older adults. Neuroimage. 2016;131:91–101.

23. Raposo N, Zanon Zotin MC, Schoemaker D, Xiong L, Fotiadis P, Charidimou A, Pasi M, Boulouis G, Schwab K, Schirmer MD, et al. Peak Width of Skeletonized Mean Diffusivity as Neuroimaging Biomarker in Cerebral Amyloid Angiopathy. AJNR Am J Neuroradiol. 2021;42:875–881.

24. McCreary CR, Beaudin AE, Subotic A, Zwiers AM, Alvarez A, Charlton A, Goodyear BG, Frayne R, Smith EE. Cross-sectional and longitudinal differences in peak skeletonized white matter mean diffusivity in cerebral amyloid angiopathy. NeuroImage: Clinical. 2020;27:102280.

25. Salthouse TA. The processing-speed theory of adult age differences in cognition. Psychological Review. 1996;103:403–428.

26. Salthouse TA. Aging and measures of processing speed. Biological Psychology. 2000;54:35–54.

27. Deary IJ, Johnson W, Starr JM. Are processing speed tasks biomarkers of cognitive aging? Psychol Aging. 2010;25:219–228.

28. Kievit RA, Davis SW, Griffiths J, Correia MM, Cam-CAN, Henson RN. A watershed model of individual differences in fluid intelligence. Neuropsychologia. 2016;91:186– 198.

29. Bollen KA. Structural equations with latent variables. Oxford, England: John Wiley & Sons; 1989.

30. Kline RB. Principles and practice of structural equation modeling, 4th ed. New York, NY, US: Guilford Press; 2016.

31. Miyake A, Friedman NP, Emerson MJ, Witzki AH, Howerter A, Wager TD. The unity and diversity of executive functions and their contributions to complex “Frontal Lobe” tasks: a latent variable analysis. Cognitive psychology. 2000;41:49–100.

32. Miyake A, Friedman NP. The Nature and Organization of Individual Differences in Executive Functions: Four General Conclusions. Current Directions in Psychological Science. 2012;21:8–14.

33. Shafto MA, Tyler LK, Dixon M, Taylor JR, Rowe JB, Cusack R, Calder AJ, Marslen-Wilson WD, Duncan J, Dalgleish T, et al. The Cambridge Centre for Ageing and Neuroscience (Cam-CAN) study protocol: a cross-sectional, lifespan, multidisciplinary examination of healthy cognitive ageing. BMC Neurol. 2014;14:204.

34. Taylor JR, Williams N, Cusack R, Auer T, Shafto MA, Dixon M, Tyler LK, Cam-CAN, Henson RN. The Cambridge Centre for Ageing and Neuroscience (Cam-CAN) data repository: Structural and functional MRI, MEG, and cognitive data from a cross-sectional adult lifespan sample. NeuroImage. 2017;144:262–269.

35. Folstein MF, Folstein SE, McHugh PR. “Mini-mental state.” Journal of Psychiatric Research. 1975;12:189–198.

36. McDonough IM, Bischof GN, Kennedy KM, Rodrigue KM, Farrell ME, Park DC. Discrepancies between fluid and crystallized ability in healthy adults: a behavioral marker of preclinical Alzheimer’s disease. Neurobiology of aging. 2016;46:68–75.

37. Kievit RA, Davis SW, Griffiths J, Correia MM, Cam-CAN, Henson RN. A watershed model of individual differences in fluid intelligence. Neuropsychologia. 2016;91:186– 198.

38. Tsvetanov KA, Henson RNA, Tyler LK, Razi A, Geerligs L, Ham TE, Rowe JB. Extrinsic and Intrinsic Brain Network Connectivity Maintains Cognition across the Lifespan Despite Accelerated Decay of Regional Brain Activation. J Neurosci. 2016;36:3115– 3126.

39. Rosseel Y. lavaan: An R Package for Structural Equation Modeling. Journal of Statistical Software. 2012;48:1–36.

40. Cusack R, Vicente-Grabovetsky A, Mitchell DJ, Wild CJ, Auer T, Linke AC, Peelle JE. Automatic analysis (aa): efficient neuroimaging workflows and parallel processing using Matlab and XML. Frontiers in neuroinformatics. 2014;8:90.

41. Winzeck S. Methods for Data Management in Multi-Centre MRI Studies and Applications to Traumatic Brain Injury. 2021 [cited 2024 Nov 29];Available from: https://www.repository.cam.ac.uk/handle/1810/323664

42. Smith SM, Jenkinson M, Johansen-Berg H, Rueckert D, Nichols TE, Mackay CE, Watkins KE, Ciccarelli O, Cader MZ, Matthews PM, et al. Tract-based spatial statistics: Voxelwise analysis of multi-subject diffusion data. NeuroImage. 2006;31:1487–1505.

43. Henriques1 RN, Henson R, Cam-CAN, Correia MM. Unique information from common diffusion MRI models about white-matter differences across the human adult lifespan. 2023 [cited 2023 Aug 3]; Available from: https://arxiv.org/abs/2306.09942

44. Cook RD. Detection of Influential Observation in Linear Regression. Technometrics. 1977;19:15–18.

45. Venables WN, Ripley BD. Modern Applied Statistics with S. Fourth Edition. New York: Springer; 2002.

46. Hayes AF. Introduction to mediation, moderation, and conditional process analysis: A regression-based approach. New York, NY, US: Guilford Press; 2013.

47. Alwin DF, Hauser RM. The Decomposition of Effects in Path Analysis. American Sociological Review. 1975;40:37.

48. Schermelleh-Engel K, Moosbrugger H, Müller H. Evaluating the Fit of Structural Equation Models: Tests of Significance and Descriptive Goodness-of-Fit Measures. Methods of Psychological Research. 2003;8:23–74.

49. Hayes AF, Scharkow M. The Relative Trustworthiness of Inferential Tests of the Indirect Effect in Statistical Mediation Analysis: Does Method Really Matter? Psychol Sci. 2013;24:1918–1927.

50. R Core Team. A Language and Environment for Statistical Computing R Foundation for Statistical Computing, Vienna, Austria [Internet]. 2020; Available from: http://www.R-project.org/

51. Levin RA, Carnegie MH, Celermajer DS. Pulse Pressure: An Emerging Therapeutic Target for Dementia. Front. Neurosci. [Internet]. 2020 [cited 2021 Feb 4];14. Available from: https://www.frontiersin.org/articles/10.3389/fnins.2020.00669/full?utm_source=fweb&utm_medium=nblog&utm_campaign=ba-sci-fnins-pulse-pressure-dementia

52. Fuhrmann D, Nesbitt D, Shafto M, Rowe JB, Price D, Gadie A, Tyler LK, Brayne C, Bullmore ET, Calder AC, et al. Strong and specific associations between cardiovascular risk factors and white matter micro- and macrostructure in healthy aging. Neurobiology of Aging. 2019;74.

53. Tsvetanov KA, Henson RNA, Rowe JB. Separating vascular and neuronal effects of age on fMRI BOLD signals. Philosophical Transactions of the Royal Society B: Biological Sciences. 2020;

54. Pinto E. Blood pressure and ageing. Postgraduate Medical Journal. 2007;83:109–114.

55. Steppan J, Barodka V, Berkowitz DE, Nyhan D. Vascular Stiffness and Increased Pulse Pressure in the Aging Cardiovascular System. Cardiology Research and Practice. 2011;2011:1–8.

56. Schaich CL, Malaver D, Chen H, Shaltout HA, Zeki Al Hazzouri A, Herrington DM, Hughes TM. Association of Heart Rate Variability With Cognitive Performance: The Multi-Ethnic Study of Atherosclerosis. JAHA. 2020;9:e013827.

57. Forte G, Favieri F, Casagrande M. Heart Rate Variability and Cognitive Function: A Systematic Review. Front Neurosci [Internet]. 2019 [cited 2021 Mar 18];13. Available from: https://www.ncbi.nlm.nih.gov/pmc/articles/PMC6637318/

58. Mahinrad S, Jukema JW, van Heemst D, Macfarlane PW, Clark EN, de Craen AJM, Sabayan B. 10-Second heart rate variability and cognitive function in old age. Neurology. 2016;86:1120–1127.

59. Tomoto T, Repshas J, Zhang R, Tarumi T. Midlife aerobic exercise and dynamic cerebral autoregulation: associations with baroreflex sensitivity and central arterial stiffness. Journal of Applied Physiology. 2021;131:1599–1612.

60. Elahi FM, Alladi S, Black SE, Claassen JAHR, DeCarli C, Hughes TM, Moonen J, Pajewski NM, Price BR, Satizabal C, et al. Clinical trials in vascular cognitive impairment following SPRINT-MIND: An international perspective. Cell Reports Medicine. 2023;4:101089.

61. The SPRINT Research Group. A Randomized Trial of Intensive versus Standard Blood-Pressure Control. N Engl J Med. 2015;373:2103–2116.

62. Kivipelto M, Mangialasche F, Ngandu T. Lifestyle interventions to prevent cognitive impairment, dementia and Alzheimer disease. Nat Rev Neurol. 2018;14:653–666.

63. Ngandu T, Lehtisalo J, Solomon A, Levälahti E, Ahtiluoto S, Antikainen R, Bäckman L, Hänninen T, Jula A, Laatikainen T, et al. A 2 year multidomain intervention of diet, exercise, cognitive training, and vascular risk monitoring versus control to prevent cognitive decline in at-risk elderly people (FINGER): a randomised controlled trial. The Lancet. 2015;385:2255–2263.

64. Maxwell SE, Cole DA, Mitchell MA. Bias in Cross-Sectional Analyses of Longitudinal Mediation: Partial and Complete Mediation Under an Autoregressive Model. Multivariate Behavioral Research. 2011;46:816–841.

65. Bleakley C, Hamilton PK, Pumb R, Harbinson M, McVeigh GE. Endothelial Function in Hypertension: Victim or Culprit? J Clin Hypertens. 2015;17:651–654.

66. Gallo G, Volpe M, Savoia C. Endothelial Dysfunction in Hypertension: Current Concepts and Clinical Implications. Front. Med. 2022;8:798958.

67. Tomoto T, Tarumi T, Zhang R. Central arterial stiffness, brain white matter hyperintensity and total brain volume across the adult lifespan. Journal of Hypertension [Internet]. 2023 [cited 2023 Mar 24];Publish Ahead of Print. Available from: https://journals.lww.com/10.1097/HJH.0000000000003404

68. Suri S, Bulte D, Chiesa ST, Ebmeier KP, Jezzard P, Rieger SW, Pitt JE, Griffanti L, Okell TW, Craig M, et al. Study Protocol: The Heart and Brain Study. Frontiers in Physiology. 2021;12:364.

69. Verhaaren BFJ, Vernooij MW, de Boer R, Hofman A, Niessen WJ, van der Lugt A, Ikram MA. High blood pressure and cerebral white matter lesion progression in the general population. Hypertension. 2013;61:1354–1359.

70. Maniega SM, Valdés Hernández MC, Clayden JD, Royle NA, Murray C, Morris Z, Aribisala BS, Gow AJ, Starr JM, Bastin ME, et al. White matter hyperintensities and normal-appearing white matter integrity in the aging brain. Neurobiol Aging. 2015;36:909–918.

71. Wilkinson GN, Rogers CE. Symbolic Description of Factorial Models for Analysis of Variance. Applied Statistics. 1973;22:392.

